# Update of transmission modelling and projections of *gambiense* human African trypanosomiasis in the Mandoul focus, Chad

**DOI:** 10.1101/2021.09.22.21263989

**Authors:** Kat S Rock, Ching-I Huang, Ronald E Crump, Paul R Bessell, Paul E Brown, Inaki Tirados, Philippe Solano, Marina Antillon, Albert Picado, Severin Mbainda, Justin Darnas, Emily H Crowley, Steve J Torr, Mallaye Peka

## Abstract

**Background:** In recent years, an integrated programme of vector control, screening and treatment of *gambiense* human African trypanosomiasis (gHAT) infections has led to a rapid decline in cases in the Mandoul disease focus of Chad. In this study, we assess whether elimination of transmission has already been achieved in the region despite low-level case reporting, quantify the role of intensified interventions in transmission reduction, and predict the trajectory of gHAT in Mandoul for the next decade under a range of control scenarios.

**Method:** We utilise human case data (2000–2019) to update a previous model of transmission of gHAT in Mandoul. We also test the updated model, which now has refined assumptions on diagnostic specificity of the current algorithm and an improved fitting method, via a data censoring approach.

**Results:** We conclude that passive detection rates have increased due to improvements in diagnostic availability in fixed health facilities since 2015, by 2.1-fold for stage 1 detection, and by 1.5-fold for stage 2. We find that whilst the diagnostic algorithm for active screening is estimated to be highly specific (99.93%, 95% CI: 99.91–99.95%), the high screening level and limited remaining infection means that some recently reported cases might be false positives, especially the ones that were not parasitologically confirmed. We also find that the focus-wide tsetse vector reduction estimated through model fitting (99.1%, 95% CI: 96.1–99.6%) is comparable to the very high reduction previously measured by the decline in catches of tsetse from monitoring traps. In line with previous results, the model suggests that transmission was likely interrupted in 2015 as a result of intensified interventions.

**Conclusions:** We recommend that additional confirmatory testing is performed in Mandoul in order that the endgame can be carefully monitored now that infection levels are so low. More specific measurement of cases would better inform when it is safe to stop active screening and vector control.

## Introduction

A new World Health Organization (WHO) roadmap [1] has set out control, elimination or eradication goals to be achieved by 2030 for 20 different neglected tropical diseases (NTDs) – a collection of mainly infectious diseases affecting some of the poorest and most marginalised populations globally. Amongst them is *gambiense* human African trypanosomiasis (gHAT), a vector-borne, parasitic infection which has a goal of elimination of transmission (EOT) by 2030 following the success indicated by decline in global disease reporting in the last two decades [2]. Formerly, this disease – caused by *Trypanosoma brucei gambiense* and transmitted by tsetse (*Glossina*) to humans – impacted 36 countries across West and Central African [3], usually resulting in death without detection and treatment of those infected. Now only 24 countries are considered endemic – at least marginal risk – and, of these, six have reported only tens of cases for the last 5 years and nine have reported single figures [2].

gHAT is a slow progressing disease with early (stage 1) infection causing relatively mild or non-specific symptoms such as headache or fever [4] with the parasite found in blood and lymph fluid. Following a period of 1–2 years [5], infection penetrates the blood-brain barrier where it causes late stage (stage 2) disease and the parasite may be found in cerebrospinal fluid (CSF). During stage 2 patients may suffer neurological symptoms including, but not limited to, neuropsychiatric symptoms, sleep disturbance, abnormal gait or movement, and eventually death [4, 6]. gHAT is not vaccine preventable nor is it currently possible to use mass distribution of drugs to treat infection. Patients will only be treated following identification of the parasite in the blood, lymph system or CSF, or in some settings as a results of strong serological evidence. Consequently there is a large diagnostic component to medical interventions and cases must be identified either via mass screening of village populations in endemic areas (active screening) or rely on fixed health care facilities, known as “passive” detection. A critical difference between passive and active screening is that all individuals are screened in active screening irrespective of their symptomatic status whilst passive screening only screens those with gHAT symptoms. Due to the non-severe clinical signs in stage 1 and the differential diagnosis with malaria, most passively detected cases are found in stage 2, at which time they could have been infected and potentially infectious for several years. More details on the differences of active and passive screening can be found elsewhere [4].

Initial screening for gHAT is performed using either the card agglutination test for trypanosomiasis (CATT) or rapid diagnostic tests (RDTs) followed by by various microscopy techniques to establish the presence of active infection. Following diagnosis it must be established whether the infection is stage 1 or stage 2 – determined by either the presence of trypanosomes or more than 5 white blood-cells per *μ*l in CSF. Up until now, this staging has been necessary for the administration of stagespecific drugs. In recent years the treatments have been intravenously-administered pentamidine for stage 1 and nifurtimox-eflornithine combination treatment (NECT) for stage 2, over the course of 10–14 days. This has changed in recent years with the introduction of fexinidazole which has obviated the requirement for staging in all but young patients and severely infected patients [6].

Detection and treatment of infected people has been the primary method to control disease and reduce transmission by reducing the time people spend infected, and thereby limiting opportunities for transmission back to tsetse vectors. However, the vector-borne nature of transmission provides other complementary options for control, including targeting the flies directly. A range of different ways to reduce tsetse populations exist, including ground and aerial spraying, insecticide-treated cattle, and insecticide-impregnated targets [7, 8].

The large reductions in cases globally between 1998 and today are broadly attributed to tools used for diagnosis and treatment of infections [2], including CATT and NECT. However, in the last decade the advances in RDTs and vector control have been accelerating progress in specific regions [9, 10, 11, 12]. Chad is one of the countries now reporting very small numbers of cases – in 2002 there was a peak of 715 cases of gHAT reported, making it the country with the fifth highest burden, but in 2019 there were just 16 cases, falling to seventh place relative to all endemic countries [13]. Reductions between 2000 and 2013 are attributed to active screening of at-risk populations and passive screening in endemic foci, however in Chad’s main focus of Mandoul, additional interventions began in 2014 and 2015 to accelerate towards zero. Vector control started in 2014 using Tiny Targets – with a measured tsetse density reduction of 99% after four months, and in 2015 the passive screening system was fortified by increasing the number of fixed health facilities with RDTs [11]. The focus of gHAT in Mandoul presents a unique opportunity for gHAT control as it is a relatively small area with a relatively small population. What is more, the tsetse habitat is a single discrete area with no scope for reinvasion or importation of vectors from other areas.

Previous modelling work suggested that it was likely that transmission was already interrupted by 2015 [11], with 62.8% (95% CI: 59–66%) of the transmission reduction due to vector control. However, in the present study we question whether this is consistent with the low-level but persistent case reporting still occurring in the focus. There are numerous other questions surrounding transmission and reporting in the Mandoul focus: (i) given there were six reported cases in 2018 and 11 in 2019, what does this tell us about underlying transmission? (ii) when can we expect to observe zero cases reported? (iii) can active screening and vector control be stopped without risking recrudescence?

In the present study we critique and refine the previous modelling work to reassess likely transmission, and update predictions until 2030 utilising a further four years of case data. New results are also available via an interactive graphic user interface (https://hatmepp.warwick.ac.uk/Mandoulfitandproject/v1/).

## Methods

### Data

To quantify the transmission and case reporting of gHAT in the Mandoul focus of Chad, we utilised the WHO HAT Atlas data from 2000–2018 [2]. Records within this data set were annually aggregated by mode of screening/detection (i.e. active and passive), and location. In a single record the number of people screened, cases identified and staging (if known) were recorded. In order to fit our model to the focus-level data we aggregated all locations with the Mandoul focus together, but retained information on the number of active (stage 1, stage 2 and stage unknown) cases, passive (stage 1, stage 2 and stage unknown) cases, and the number of people actively screened in each year. To supplement these data, we also used 2019 case and screening data provided by PNLTHA-Chad. The geographical location and approximate extent of the Mandoul focus and other foci in Chad are presented in supplemental Figure 1.

In Chad, case identification follows an algorithm involving serological screening using CATT (in vehicle based active screening) and RDT (in passive screening and motorcycle based active screening). Confirmation of the presence of circulating parasites is by various microscopy techniques, supplemented in 2017 by the highly-sensitive mini anion exchange centrifugation technique (mAECT) technique. However, in the absence of positive microscopy, cases are also confirmed by clinical signs and positive results by CATT on diluted serum. For some suspects additional (but not final) diagnostic data is obtained using loop-mediated isothermal amplification (LAMP).

### The gHAT model

Mechanistic transmission modelling is one way to represent dynamic changes in the spread of an infection over time. It can account for variable interventions (in particular active screening), or introduction of a new strategy (such as improved passive screening and vector control). Once calibrated to data, models can be used to predict what we might expect to happen in the future if the current intervention strategy is continued, or if changes are made.

The Warwick gHAT model has been developed over the past five years to represent the known biology of transmission between humans and tsetse (and possibly non-human animals), and capture key gHAT intervention strategies used in both the Democratic Republic of Congo and Chad. Since its original development [14] several modifications have been made based on continual fitting to different longitudinal data sets, followed by assessment and refinement [15, 16, 11, 17]. In the present study we specifically discuss the evolution of the Warwick gHAT model since it was original fitted to data from the Mandoul focus [11] and take steps to update the previous fits and provide new projections for the future under different strategies.

### Assessment and update of model fits

In order to quantitatively assess previous model projections and refine them we take three steps.

#### Step 1: Previous model projections corrected for new active screening

First we use the previously developed model code and posterior parameterisation from fitting to data from 2000-2013 [11] and use the active screening numbers from 2014–2019 to update projections for those six years. In Mahamat *et al*., screening numbers for 2014 and 2015 were known, but numbers for 2016–2019 were not. Large differences between assumed screening levels and actual screening levels can have a substantial impact on model projections - if there is no screening there would be no active cases, whereas there could be a high number of cases if large proportions of the population were screened. In Mahamat *et al*., it was assumed that the screening numbers for 2016–2019 would be 27,265 each year (the same as in 2015), whereas new data from the WHO HAT Atlas and PNLTHA are somewhat lower – 22,0071, 18,144, 18,083 and 12,640 for each of the years respectively. This first step enables us to compare new data with old model predictions (adjusted for the correct screening numbers). In line with the previous results, we utilised ensemble results from four different model variants, two of which include animal reservoirs (see SI for more details on the previous model).

Following fitting in Mahamat *et al*. [11], counterfactual scenarios were run for 2014–2019 to estimate the impact of intensified interventions. For counterfactual scenarios considered here, we simulated what would have been expected if (a) neither vector control nor improved passive screening had been introduced in 2014 and 2015, (b) if the only change was improved passive screening, and (c) if the only change was vector control. These three scenarios were compared to the “actual” strategy (with both vector control and improved passive screening) in order to assess the relative impact of the different intervention types in reducing transmission.

#### Step 2: New model fit for 2000–2013

Since the publication of Mahamat *et al*. [11], there have been several refinements to the underlying gHAT model used. Amongst these, the model is now able to capture greater variance in observed case reporting (overdispersion) which was originally developed for Model W in Castano *et al*. [15], and the model fitting procedure was improved by using informative priors which take previous biological beliefs about the parameterisation into account (e.g. the high-risk group in the population is likely a similar proportion to the proportion of working age men and *R*_0_ is probably only slightly above 1) [17]. Furthermore modelling of the improvement to passive screening (used from 2015 onwards to simulate increased RDT usage in Mandoul) has a slightly different formulation: from a discrete jump in detection rates to the use of a steep logisitic function, although the consequence of this change is expected to be minimal (see SI). In the previous model, underreporting in passive screening was assumed to occur with both stage 1 and 2 infections, however subsequent updates now assume that true stage 1 infection either leads to reported cases or progression to stage 2. Stage 2 infections can still lead to underreported deaths, with the parameter *u* dictating the (passive) reporting probability of stage 2 infections not picked up by active screening.

The specificity of the diagnostic algorithm in previous modelling was assumed to be 100%, whereas PNLTHA-Chad currently allow for treatment based on either a serological or parasitological diagnosis. Patients who are only positive by serology must remain positive after a repeat CATT test is performed with 1:8 dilution, rather than only on the initial CATT on whole blood, and these patients are termed “strongly” seropositive cases. The inclusion of these strongly seropositive individuals as cases has the potential to miss fewer remaining infections who might otherwise be false negative in parasitology, and therefore treat these people and shrink the remaining infectious reservoir. However, it also risks over-diagnosis of cases (false positives) due to the high number of people screened per year, even with a high estimated specificity of over 99% for an algorithm including CATT 1:16 case confirmation (99.1%: 95% CI: 97.7–99.7% [18], 99.36%: 95% CI: 99.22–99.48%[19], and *>*99.6% [20]). As infection continues to decline in Mandoul – and globally – the positive predictive value of tests is reduced and eventually false positives may outnumber true positives without parasitological confirmation or laboratory-based follow-up using molecular diagnostic tools such as Trypanolysis [21] which are extremely specific. The new model, therefore, fits specificity to the data using the literature estimates as a prior. Our initial assumption is that any false positives would be assigned in the staged data as Stage 1.

Using the new model code and fitting procedure, our second step is to fit the model again to 2000–2013, making projections for 2014–2019 using the same assumptions about improvements to VC and PS in 2014 and 2015 respectively, specifically a 99% tsetse reduction after four months and a doubling of both the stage 1 and stage 2 passive detection rates. By comparing step 1 and 2 we can see the impact of model improvements, but not improvements due to more data availability. As with previous fitting, we fitted the same eight different model variants.

#### Step 3: New model fit for 2000–2019

Finally our third step is to fit the full data set including 2014–2019. This includes inferring (i) the stage 1 and stage 2 PS improvement from 2015 and (ii) the overall reduction of tsetse in the whole epidemiological focus (rather than using the measured tsetse density reduction in the Mandoul study area). Estimating the tsetse reduction using the model fitting, rather than substituting the value measured in the field allows us to test whether there is agreement between model outputs based on human case data and entomological dynamics observed in the study region. Any differences in these two could indicate that infection is occurring outside of the area under control by Tiny Targets. This third step results in the inference of three extra parameters, parameters 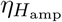 and 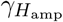 which dictate the increased amplitude of the passive detection rate in stage 1 and stage 2 respectively, and the probability of a tsetse receiving a lethal dose of insecticide during the host-seeking phase of its feeding cycle, *p*_targetdie_, for vector control. See the SI for more model details.

Based on preliminary results which assumed false positives must be stage 1, we relaxed this assumption and allowed false positives to be reported as either stage 1 or stage 2 with a fitted probability.

As in the original study, counterfactual scenarios were run for 2014–2019 to estimate the impact of intensified interventions on transmission and reporting. Since the actual strategy for this time period is now fitted rather than projected, this should provide a more robust estimate than reported previously.

Table 1 summarises assumptions for the three model fits described above.

**Table 1.** Comparison of the fits of previous and new models.

|  | Previous model | New model fit for 2000–2013 | New model fit for 2000–2019 |
| --- | --- | --- | --- |
| **Active screening** |  |  |  |
| Specificity | 100% | Estimated from data | Estimated from data |
| False positives (FP) | No FP | FP in stage 1 only | FP in either stage |
| **Passive screening** |  |  |  |
| Improvement rate | Doubling of stage 1 and stage 2 PS detection rates from 2015 | Doubling of stage 1 and stage 2 PS detection rates from 2015 | Stage 1 and stage 2 PS detection rates from 2015 were estimated from the data |
| Underreporting | Both stages | Stage 2 only | Stage 2 only |
| **Vector control** |  |  |  |
| Reduction after 4 months | 99% | 99% | Total reduction estimated from the data |

### Projections and cessation

Using the 2000–2019 model fit we also make projections until 2050 under five strategies (see Table 2), comprising of continued AS at either mean (during 2015–2019) or maximum coverage (during 2000–2019, MaxAS) with imperfect (*<*100%, fitted) and perfect (=100%) test specificity in conjunction with or without continuation of vector control from 2021. In all scenarios passive screening was assumed to remain at present levels. Two MaxAS+VC strategies are presented in the SI. Due to resource limitations, including costs and effort to implement such programmes, we also examined a cessation criterion for active screening and vector control following three years of zero case detections to consider the impact that stopping activities based on this measure could have (see SI for sensitivity analysis on cessation criteria). The earliest cessation year was assumed to be 2021. Reactive AS takes place when any passive detections occur after the cessation criterion is met. The coverage of AS depends on its projection strategy. The duration of reactive AS depends on case reporting, such that reactive AS stops again when no case (either active or passive) is found in that year.

**Table 2.**
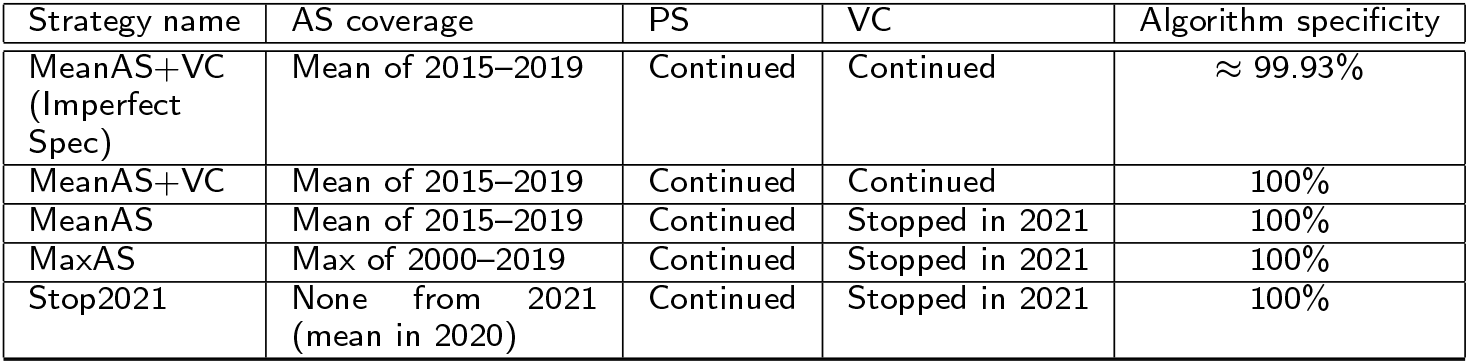
Future strategies (2020 onwards) considered in the present study. AS = active screening, PS = passive screening, VC = vector control.

| Strategy name | AS coverage | PS | VC | Algorithm specificity |
| --- | --- | --- | --- | --- |
| MeanAS+VC<br>(Imperfect Spec) | Mean of 2015–2019 | Continued | Continued | $\approx 99.93\%$ |
| MeanAS+VC | Mean of 2015–2019 | Continued | Continued | 100% |
| MeanAS | Mean of 2015–2019 | Continued | Stopped in 2021 | 100% |
| MaxAS | Max of 2000–2019 | Continued | Stopped in 2021 | 100% |
| Stop2021 | None from 2021<br>(mean in 2020) | Continued | Stopped in 2021 | 100% |

## Results

### Assessment of model fits

The previous model predicted that there would be a median of zero cases by 2017 in active screening and by 2018 in passive screening. Newly available data from 2017– 2019 are very low, but cases were still reported, indicating that our previous model overestimated the impact of the strategy on case reporting from 2014. Correcting for the real screening coverage had little impact on our predicted cases for this time period, due to the very limited remaining infection estimated by the model.

The updated model fit for the same fitted time period as before (2000–2013) better captures variation in case observations during the training period, attributed to the inclusion of overdispersion parameters in the updated model and improvement in the automated MCMC algorithm. It also matches more closely with the 2014–2019 data despite having no more information than the previous model for fitting and making the same assumptions on the vector control reduction and passive detection rate improvement. Again, the overdispersion in the updated model leads to wider prediction intervals, more reflective of our uncertainty. In addition, the model now accounts for imperfect specificity in the diagnostic algorithm; not only does that allow for the potential of some false positives in 2017–2019, but it also impacts the model fit for 2000–2013. Median active detection predictions increased to 26, 26 and 18 for 2017–2019 compared to zero for the previous model with new screening data, and median passive detection predictions increased to 7, 2 and 0 from 1, 0 and 0 for the same time frame. The updated fit is therefore more in line with real data from these years (actively reported cases were 15, 3 and 10, and passively reported cases were 7, 3 and 1 for 2017–2019). Furthermore the enlarged prediction intervals for case reporting in the updated model now cover all the reported data for the prediction period except passive cases in 2015 and active cases in 2018.

The number of new infections per year (a measure of transmission) is a quantity which is very important with respect to the 2030 elimination goal, but ultimately is very difficult to measure directly in the field. The mechanistic model types used in this study and Mahamat *et al* [11] can provide an estimate of transmission levels that would be required in order to generate the observed case reporting in active and passive detection. Factors such as underreporting and accuracy of diagnostic screening tests can impact the inferred level of transmission. Model estimates for transmission can be seen SI Figures 5, 6 and 7, and demonstrate that whilst the updated model fitted to the full data set infers slightly lower transmission in the early 2000s, the decrease between 2000 and 2013 follows the same trend (72% median reduction in the previous model and 68% median reduction in the updated model fitted to the 2000–2019 data). Imperfect specificity of the diagnostic algorithm captured in the updated model (but not the original) is one explanation for lower new infections; if some reported cases were false positive then the infections would be expected to be less than if all case reporting is true positives. The reporting parameter, *u*, can also impact inference of new infections. In the updated model fit (2000–2013), *u* was found to be around 20–33%, slightly higher than previously estimated using the same data (*u* was 19.1% median (95% CI: 16.8–23.6%)). Analogous to imperfect specificity, lower underreporting of passive cases means that for the same number of new infections there would be more passive cases, or conversely, for the same number of passive cases there are actually fewer infections.

Despite the good agreement between aggregated active cases in the data and model (2000–2013) fit for the projection period, the model predicted that almost all reported cases would be false-positive from 2015 onward, whereas there are similar case detections in stage 1 as in stage 2 in the data. Therefore in the full data set fit (2000–2019) we decided to allow for false positive detections to be assigned to either stage, rather than only stage 1. Using the data from 2000–2013, the reported cases were too high to be able to estimate this trend as most would be expected to be true positive cases.

The updated model fitted to the full data set (2000–2019) enabled us to estimate the reduction in vectors in 2014 and the improvement to passive detection rates from 2015. Through our model calibration we found that the focus-wide reduction of tsetse (after four months) was estimated to be 99.1% (95% CI: 96.1–99.6%), around our 99% assumed estimate based on the catch of tsetse from monitoring traps in the intervention area. For passive detection we estimated that stage 1 rate (*η*_*H*_) increased to 2.12 times previous levels (95% CI 1.19–4.06) and stage 2 rate (*γ*_*H*_) was 1.52 times previous levels (95% CI 1.04–8.60). This suggests that the doubling assumed for stage 1 passive detection in the previous model was quite reasonable, although it was slightly high for stage 2.

Using this full fit we estimated that the active screening algorithm had a specificity of 0.9993 (95% CI: 0.9991–0.9995) and that false positive cases were slightly more likely to be diagnosed as stage 2 rather than stage 1 (0.61 (95% CI: 0.46– 0.74)). Using the information directly from the data collected during 2016–2019 we can see that, proportionally, many of the cases reported in active screening had positive serology but were not confirmed through parasitology (see SI Figure 10). Full posterior distributions for these and other model parameters can be found in the SI (Table 8 and Figure 3).

### Counterfactual scenarios

By simulating counterfactual scenarios (CFSs) without one or both intensified interventions occurring in 2014 and 2015, the model can be used to estimate case reporting and transmission that would have likely occurred if previous active screening and baseline passive screening had continued. Table 3 shows the reduction in transmission for the first two-year period and first six-year period following implementation of intensified interventions. It also provides the percentage attributions of the basic strategy, enhanced passive screening and vector control in achieving the reductions. Notably, vector control is computed to have had the most impact on transmission amongst all the interventions for both time periods, however both intensified interventions were inferred to have substantial impact over the six-year period (23.6% for enhanced passive screening and 34.7% for vector control). Even without new interventions, active and passive screening alone would have likely reduced transmission, but not enough to interrupt transmission by the target year of 2030 (see SI Figure 8); under the strategy including enhanced passive screening but no vector control, there was a prediction of a 6.6% probability of EOT when performed in conjunction with mean active screening. With the conducted intervention the estimated year of EOT was 2015 (95% PI: 2015–2015).

**Table 3.**
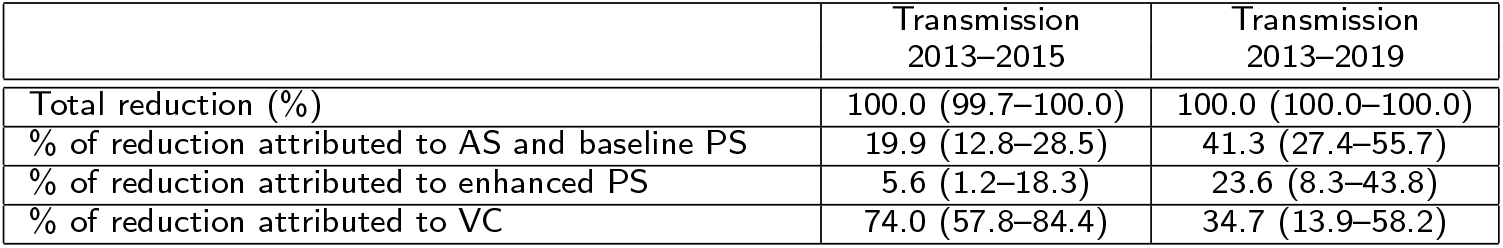
Estimated percentage reduction in transmission by intervention since intensified strategy began. Attributions to each strategy are based on counterfactual strategy simulations. Medians are given with 95% credible intervals in brackets. AS=active screening, PS=passive screening and VC=vector control.

### Evidence for alternative model structures: the evidence for and against an animal reservoir

We found that there was most support for model variants 4 and 5 - both of which have high-/low-risk structure for the human population, but do not have animal reservoirs (see SI for more details). Less than 0.1% of the ensemble model was made up of simulations from Models 7 and 8 (equivalent to Models 4 and 5 but including transmission to and from animals). Even taking these models alone, fitting indicates it is unlikely that animals constitute a maintenance reservoir, with 39% and 24% of simulations having human only maintenance of infection in Models 7 and 8 respectively, and the remainder requiring both transmission from humans and non-human animals.

### Projections to 2030

Figure 2 shows the ensemble model prediction (using the updated 2000–2019 fit) under five different strategies including three with continuation of the mean level of active screening (2015–2019), one under the maximum coverage observed during 2000–2019, and one with no active screening from 2021. In the baseline case we assume that specificity remains imperfect in active screening (99.93%), however in all other scenarios we assume that additional testing in the active screening algorithm increases specificity to 100% from 2021. We simulate cessation of active screening following three years of no case detections in Figure 2, however other cessation criteria yield virtually identical results.

**Figure 1.**
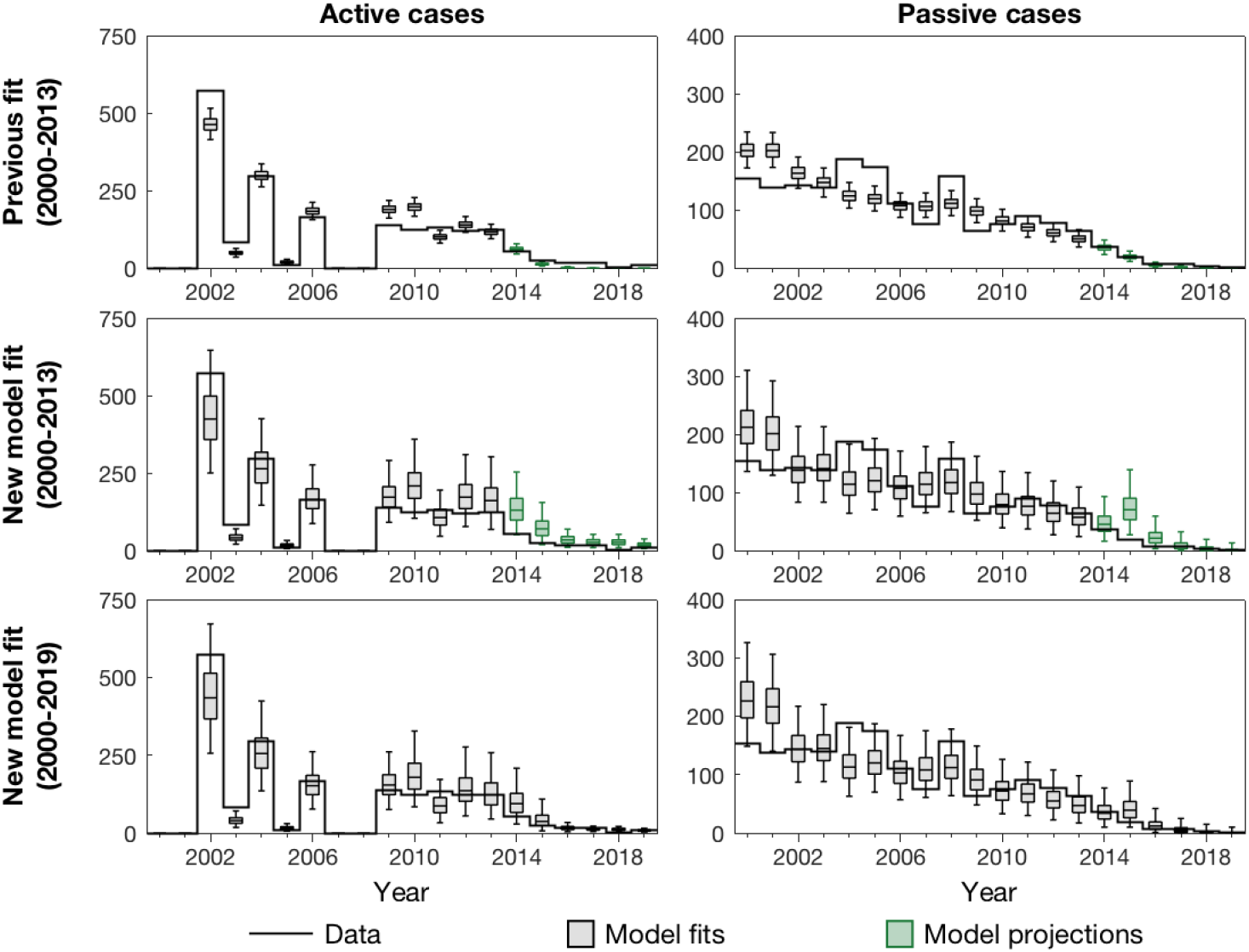
Comparison of previous and new model outputs. This figure panel shows the results of fitting to case data during (A) 2000–2013 using the previous model, (B) 2000–2013 using the new model, and (C) 2000–2019 using the new model. The solid black lines show the case data. Grey box and whiskers indicate years of model fits (median for centre line and 95% credible intervals for whiskers) and green box and whiskers denote model projections based on known active screening coverage.

**Figure 2.**
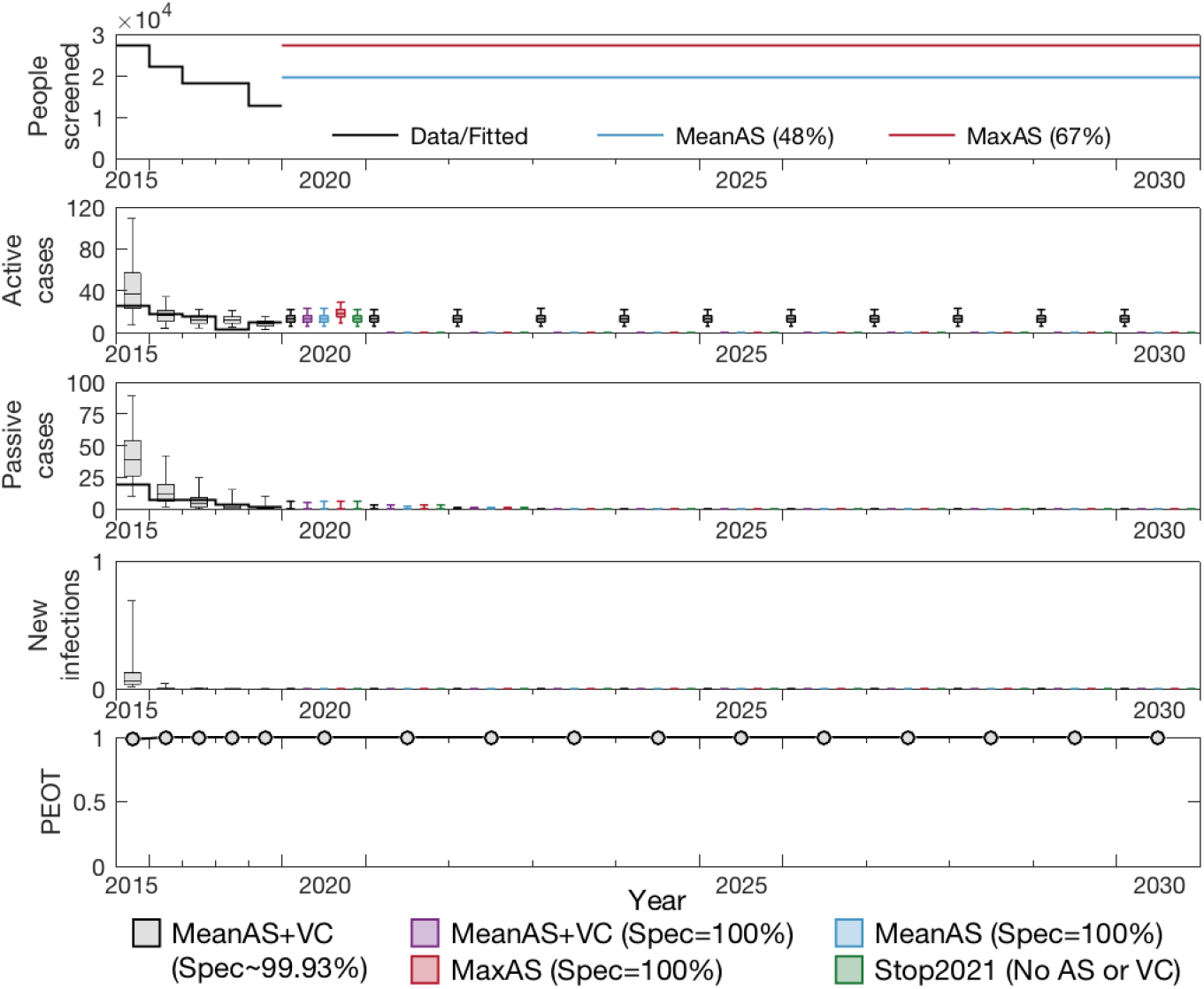
Projections to 2030. The ensemble model fitted to data during 2000–2019 was used to make projections under five different strategies. The baseline strategy, MeanAS+VC with imperfect specificity (∼ 99.93%), is denoted by grey boxes. With specificity improved to 100% from 2021, the strategy MeanAS+VC is denoted by purple boxes. Blue and red boxes are MeanAS and MaxAS strategies with AS screening specificity switching to 100% and stopping VC from 2021. Finally, Stop 2021 under which both AS and VC stop in 2021 is shown by the green boxes. All simulations assume PS remains at the level as estimated for 2019 and continues indefinitely. The top panel shows the level of AS assumed in the different projections, the second row shows the active case predictions, the third shows the passive case predictions and the forth shows the expected amount of new infections. The bottom row shows the probability of EOT for each year.

Notably, under all strategies we predict that elimination of transmission has already occurred and will not resume even after cessation of vertical interventions. In these simulations the extreme suppression of the fly population does not recover, however even if there were very rapid bounceback of tsetse through reinvasion or other means, only 2% of our model simulations (with 90% adult tsetse reintroduction in 2021) saw resurgence of infection in humans where transmission occurred beyond 2030 as a result. Our more modest reintroduction scenarios (10% and 50% in 2021) found 0% and 1% resurgence probability respectively (see SI Figure 9).

Interactive results for both the CFSs and the projections from 2020 can be found through our graphical user interface (https://hatmepp.warwick.ac.uk/Mandoulfitandproject/v1/), which shows the ensemble model results for both sets of model outputs.

## Discussion

This modelling study has critically examined previous modelling work to assess its predictive ability and to refine the results presented before [11]. Overall, we have found that the model predictions were reasonable, however slightly underestimated active case reporting in the late 2010s. A combination of model improvements and new data have allowed us to examine how predictions could be updated using refined methodology alone, and what additional information could be learnt from new data. Our updated model estimates the same elimination of transmission year as before (2015) and the previous and updated models attribute transmission reductions in quantitatively similar proportions to the different intervention activities. This modelling update brings together quantitative validation with discussions with those familiar with on-the-ground implementation, thus representing an important step for models aimed at providing policy recommendations. The present study adheres to the policy-relevant items for reporting models in epidemiology of neglected tropical diseases (NTD-PRIME) criteria, which map good modelling practice [22] (see SI for more details). Our graphical user interface also provides a more interactive way to view our new results https://hatmepp.warwick.ac.uk/Mandoulfitandproject/v1/.

Despite our updated model modification to better reflect observed cases (e.g. overdispersion in sampling and the possibility of false positive reporting), there are still elements of the biology which are not captured, including no possibility for asymptomatic, self-curing human infections [23] and no imported cases from other regions. We use a deterministic model which captures the average dynamics rather than a stochastic model, although work using both deterministic and stochastic gHAT models in other regions [24, 25, 26, 27] indicates that we would expect the dynamics to be very similar. Future work regarding stochastic reinvasion of infection via human imports (such as from other extant foci in Chad or over the nearby border with the Central African Republic) would provide insights on post-elimination risks for Mandoul, but is beyond the scope of the present study.

This study examined a variety of plausible future intervention strategies for Mandoul, however passive screening was assumed to remain intact at current operating levels in all of them. Reduction in the capacity or coverage to find and treat gHAT-infected people could have deleterious impact on the focus which otherwise anticipates reaching zero infected people in the next few years. Another concern is the possible effect of other infectious disease outbreaks on gHAT. The most notable example is in Guinea, where the West African Ebola outbreak in 2014–2016 had a large impact on the national control programme’s active and passive screening activities and is likely to have increased morbidity and mortality from this disease [28, 29]. Whilst there was some delay to active screening activities for gHAT in Chad in 2020 due to the COVID-19 pandemic, screening was able to resume later in the year yielding only a small impact on total annual coverage compared to recent years. Other factors which could lead to reduction in passive screening capacity include lack of funding, motivation, or dedicated human resources especially after several years of zero case detections. Horizontal integration within the health system could be vital for post-elimination monitoring and signs of resurgence.

Ongoing work brings together transmission modelling and costs in a health economic framework in order to provide assessment of the expected total costs and cost-effectiveness of possible future strategies in the focus.

## Conclusion

In this study we used a mathematical transmission model of gHAT to update previous predictions for the pathway to elimination in the Mandoul focus of Chad. This present study served several purposes including critiquing previous modelling work, demonstrating the improvements made to the updated model using a censoring validation step, and using new years’ data to further improve the updated model fit. Through fitting the tsetse reduction parameter and passive screening improvement parameters, we find that our previous assumption of a doubling stage 1 passive detection rate appears appropriate (95% CI: 1.19–4.06), although the stage 2 passive detection rate is unlikely to have increased substantially. The model fits inferred that the tsetse reduction after four months of initiating deployment was in good agreement with that measured by tsetse density in the intervention area – 99.1% (96.1–99.6%) via model inference compared to 99% via tsetse density – and this high level of tsetse control contributed to 74.0% (57.8%–84.4%) of the transmission reduction between 2013–2015, and indicated that the year of EOT was 2015 (95% CI: 2015–2015).

Projections suggest that we expect to have zero passive case reporting before 2023, although active case detection would continue due to imperfect specificity of the current diagnostic algorithm, and therefore additional confirmatory testing (such as mAECT, LAMP or Trypanolysis) ought to be considered..

It appears that cessation of vertical interventions (active screening and vector control) would be unlikely to result in a resurgence of infection if there are no or few importations of gHAT to the region, however continued surveillance will be important to have assurance that elimination of transmission has been met.

## Data Availability

Data cannot be shared publicly because they were aggregated from the World Health Organisation's HAT Atlas which is under the stewardship of the WHO. Data are available from the WHO (contact or visit https://www.who.int/trypanosomiasis_african/country/foci_AFRO/en/) for researchers who meet the
criteria for access. Model code and outputs produced from this study are available through Open Science Framework https://osf.io/rak9d/.

https://hatmepp.warwick.ac.uk/Mandoulfitandproject/v1/

## Author’s contributions

- Conceptualisation KSR
- Methodology KSR, REC, CH
- Software KSR, REC, CH, PEB
- Validation
- Formal Analysis KSR, CH
- Investigation KSR
- Data Curation KSR, PRB, AP, JD, SM, MP
- Writing - Original Draft KSR, CH
- Writing Review and editing KSR, CH, REC, PEB, SJT, PS, PRB, IT, SM, MA, JD, AP, EHC
- Visualisation CH PEB KSR
- Supervision
- Project administration KSR, EHC
- Funding acquisition KSR

## Ethics approval and consent to participate

As this study was a secondary analysis of aggregated programme data which was not personally identifiable, ethics approval was not required.

## Acknowledgements

The authors thank PNLTHA-Chad for original data collection and WHO for data access (in the framework of the WHO HAT Atlas [2]).

## Availability of data and materials

Data cannot be shared publicly because they were aggregated from the World Health Organisation’s HAT Atlas which is under the stewardship of the WHO. Data are available from the WHO (contact or visit https://www.who.int/trypanosomiasis_african/country/foci_AFRO/en/) for researchers who meet the criteria for access. Model code and outputs produced from this study are available through Open Science Framework https://osf.io/rak9d/.

## Additional Files

### Additional file 1 — Supplementary information

This pdf file contains more details of the model used in this analysis, and provides additional results including posterior parameter plots, model evidence for animal reservoirs, time series plots, and counterfactual scenarios.

### Additional file 2 — Graphical User Interface

The companion graphic user interface (GUI) for this study can be found at: https://hatmepp.warwick.ac.uk/Mandoulfitandproject/v1/.

### Additional file 3 — Open Science Framework

All model code for the present study can be found at: https://osf.io/rak9d/.

