## Supplemental Text for "Update of transmission modelling and projections of *gambiense* human African trypanosomiasis in the Mandoul focus, Chad"

RESEARCH

#### Supplementary Information

#### 1 Location of study area

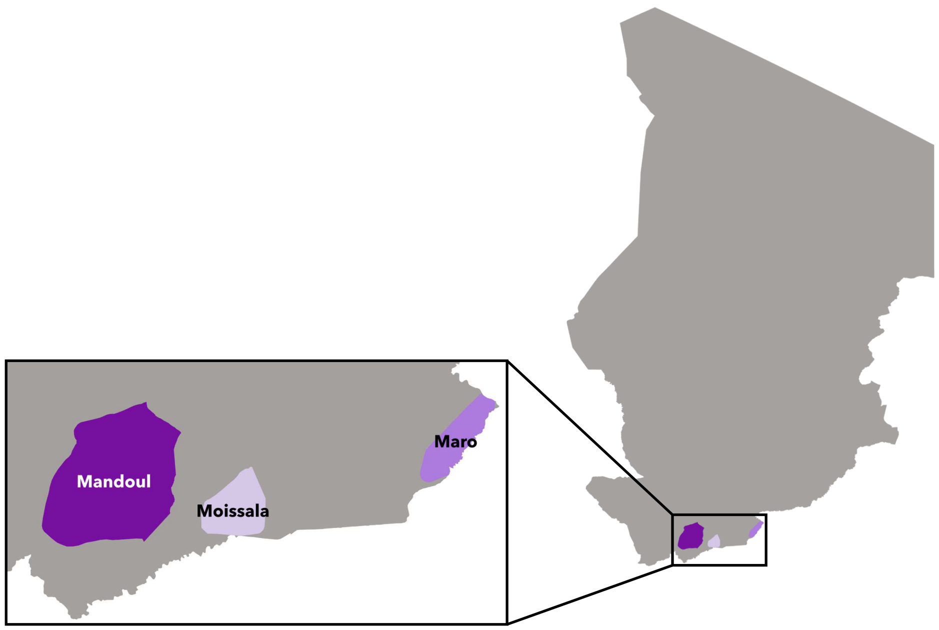


**Figure 1 Location of the Mandoul focus.** All extant gHAT foci in Chad are located along the southern border near neighbouring Central African Republic. The exact extent of the area of transmission for Mandoul is hard to precisely define, however almost all of the gHAT cases labelled as Mandoul in the WHO HAT Atlas have been geolocated to the geographic region indicated as Mandoul in dark purple.

#### 2 gHAT infection model

##### 2.1 Model variants

The gHAT model equations are given below (4) and correspond with Fig 2. In this study, eight different model variants are used, as per our previous modelling study for Mandoul [1]. Elsewhere, model variant "Model 4" has been used extensively as there is a good match to data [2, 3, 4, 5, 6].

| Model | Random participation | | Non-participation | | Animals |
|---|---|---|---|---|---|
| | Low-risk | High-risk | Low-risk | High-risk | |
| 1 | X | | | | |
| 2 | X | X | | | |
| 3 | X | | X | | |
| 4 | X | | | X | |
| 5 | X | X | X | X | |
| 6 | X | | | | X |
| 7 | X | | | X | X |
| 8 | X | X | X | X | X |

**Table 1** Different model structures under consideration

In the present study human hosts are assumed to be either at low-risk and randomly participate in active screening (subscript $H1$), at high-risk and randomly participate in active screening (subscript $H2$), at low-risk and never participate in active screening (subscript $H3$) or at high-risk and never participate in active screening (subscript $H4$). In Models 1–5 tsetse bites are assumed to be taken on humans or non-reservoir animals. However, the non-reservoir animal species are not explicitly modelled. In Models 6–8 animals capable of acquiring infection from and transmitting infection to tsetse are also included.

The model is parameterised with a combination of fixed and fitted parameters. Fixed parameters (see Table 2) generally correspond to assumed biological values that are unlikely to vary across space or are known for Chad, such as the human mortality rate, tsetse bite rate and stage 1 to stage 2 disease progression in humans. Fitted parameters (see Tables 3–6) are those which are likely to be correlated with region or are unknown, including the tsetse-to-human relative density, the proportion of the population who are at high-risk of exposure to tsetse, the reporting rate (linked to access to health facilities with gHAT testing capacity), and parameters linked to the time to passive detection.

#### 2.2 Modelling vector control

The function which describes the probability of both contacting a and dying is time dependent (days) from when the targets were placed:

$$f_T(t) = p_{\text{targetdie}} \left(1 - \frac{1}{1 + \exp(-0.068(\text{mod}(t, 365) - 127.75))}\right) \tag{1}$$

and $p_{\text{targetdie}}$ is chosen or fitted such that the tsetse population after a four month period reflects the observed/assumed percentage reduction. A value of $p_{\text{targetdie}} = 0.4171$ would yield a 99% reduction after 4 months and is approximately equivalent to an additional daily mortality of 0.14. As the present study focuses on Chad with annual deployment we assume that targets are deployed once per year, rather than twice a year which is the standard in DRC and therefore a slightly different form of the equation is used [6].

#### 2.3 Modelling improvements in passive screening

Following modelling work for fitting to gHAT case data in DRC [6], for simulations in this study, we assumed that prior to 1998 there was limited passive case detection, which would not detect stage 1 cases ($\eta_H^{\text{pre}} = 0$) and have a slower time to detection for stage 2 ($\gamma_H^{\text{pre}} = b_{\gamma_H^{\text{pre}}} \times \gamma_H^{\text{post}}$ where $b_{\gamma_H^{\text{pre}}} = [0, 1]$). In 1998 we assume that the

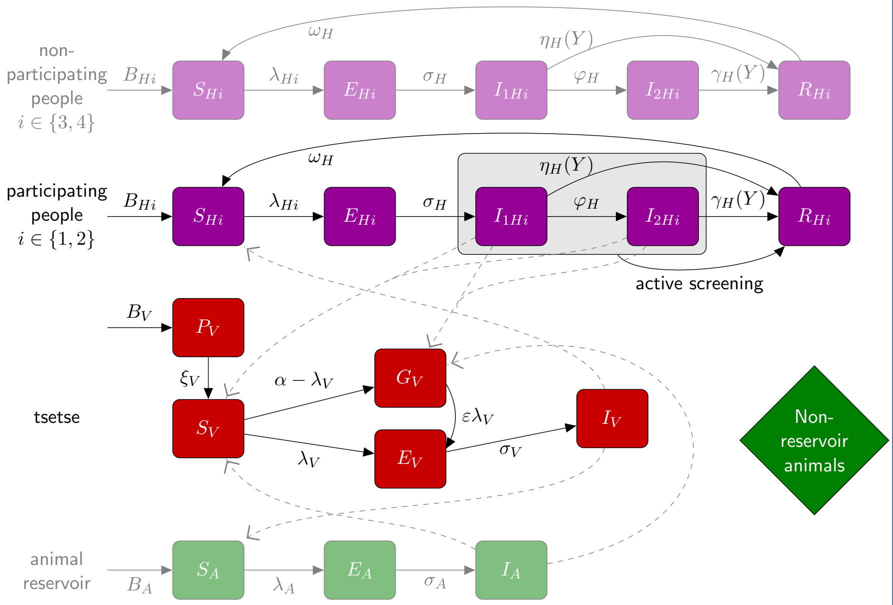


**Figure 2 Schematic of gHAT model (and its variants).** The core components which feature in every model variant (M1–M8) are the human population group $i = 1$ (low-risk and participating in active screening), tsetse and non-reservoir animals. Human populations are partitioned into infection status - susceptible ($S_{Hi}$), exposed but not infectious ($E_{Hi}$), infected in stage 1 ($I_{1Hi}$), infected in stage 2 ($I_{2Hi}$ and recovering following treatment and before returning to susceptible ($R_{Hi}$). Tsetse classes include the pupal stage ($P_V$), newly emerged and unfed/teneral flies ($S_V$), non-teneral and uninfected flies which have reduced susceptibility to infection ($G_V$), exposed and incubating flies ($E_V$) and infected flies ($I_V$). Non-reservoir animals do not confer infection but do account of a proportion of tsetse blood feeding. In model variants 6–8 there is a reservoir animal population (faded green). Other models have various combinations of participating and non-participating humans and different risk of exposure to tsetse (reflected in the force of infection parameter $\lambda_{Hi}$ which is $r > 1$ times greater for high-risk groups) shown in faded purple. This diagram is adapted from Crump *et al.* [6] and Mahamat *et al.* [1] under a creative commons licence.

introduction of the card agglutination test for trypanosomes (CATT) enabled better diagnosis and stage 1 and 2 rates were increased to $\eta_H^{\text{post}}$ and $\gamma_H^{\text{post}}$ respectively.

To simulate the improvement to passive screening in 2015, following an intervention by PNLTHA and FIND, we used logistic functions to describe improvements to both the passive stage 1 and stage 2 detection rates in each year:

$$\eta_H(Y) = \eta_H^{\text{post}} \left[1 + \frac{\eta_{H_{\text{amp}}}}{1 + \exp\left(-d_{\text{steep}}(Y - d_{\text{change}})\right)}\right] \tag{2}$$

$$\gamma_H(Y) = \gamma_H^{\text{post}} \left[1 + \frac{\gamma_{H_{\text{amp}}}}{1 + \exp\left(-d_{\text{steep}}(Y - d_{\text{change}})\right)}\right] \tag{3}$$

As there was a rapid roll out of improvements we selected a value of $d_{\text{steep}} = 10$ which pushes this logistic form to be virtually a step function. This is different to the shallower $d_{\mathrm{steep}} \approx 1$ value estimated in Bandundu province, DRC, where it is thought that more gradual improvements occurred over several years [6]. We fixed the $d_{\mathrm{change}}$ parameter to be 2014.5, which corresponds to the rate jumping up from 2015 onwards, when the intervention was first implemented.

For the updated model fit to 2000–2013 data we fixed the amplitude of both functions to be equal to 1, resulting in a doubling of detection rates from 2015 onwards. In the 2000–2019 fit we allowed the amplitude parameters to be fitted independently.

$$
\text{Humans}\begin{cases}
\dfrac{\mathrm{d}S_{Hi}}{\mathrm{d}t} &= \mu_H N_{Hi} + \omega_H R_{Hi} - \alpha m_{\text{eff}} f_i \dfrac{S_{Hi}}{N_{Hi}} I_V - \mu_H S_{Hi} \\
\dfrac{\mathrm{d}E_{Hi}}{\mathrm{d}t} &= \alpha m_{\text{eff}} f_i \dfrac{S_{Hi}}{N_{Hi}} I_V - (\sigma_H + \mu_H) E_{Hi} \\
\dfrac{\mathrm{d}I_{1Hi}}{\mathrm{d}t} &= \sigma_H E_{Hi} - (\varphi_H + \eta_H(Y) + \mu_H) I_{1Hi} \\
\dfrac{\mathrm{d}I_{2Hi}}{\mathrm{d}t} &= \varphi_H I_{1Hi} - (\gamma_H(Y) + \mu_H) I_{2Hi} \\
\dfrac{\mathrm{d}R_{Hi}}{\mathrm{d}t} &= \eta_H(Y) I_{1Hi} + \gamma_H(Y) I_{2Hi} - (\omega_H + \mu_H) R_{Hi}
\end{cases}
$$

$$
\text{Animals}\begin{cases}
\dfrac{\mathrm{d}S_A}{\mathrm{d}t} &= \mu_A N_A - \alpha m_{\text{eff}} f_A \dfrac{S_A}{N_A} I_V - \mu_A S_A \\
\dfrac{\mathrm{d}E_A}{\mathrm{d}t} &= \alpha m_{\text{eff}} f_A \dfrac{S_A}{N_A} I_V - (\sigma_A + \mu_A) E_A \\
\dfrac{\mathrm{d}I_A}{\mathrm{d}t} &= \sigma_A E_A - \mu_A I_A
\end{cases}
$$

$$
\text{Tsetse}\begin{cases}
\dfrac{\mathrm{d}P_V}{\mathrm{d}t} &= B_V N_H - (\xi_V + \frac{P_V}{K}) P_V \\
\dfrac{\mathrm{d}S_V}{\mathrm{d}t} &= \xi_V P(\text{survive pupal stage}) P_V - \alpha S_V - \mu_V S_V \\
\dfrac{\mathrm{d}E_{1V}}{\mathrm{d}t} &= \alpha\big(1 - f_T(t)\big) p_V \big( \sum_i f_i \dfrac{(I_{1Hi} + I_{2Hi})}{N_{Hi}} + f_A \dfrac{I_A}{N_A} \big)(S_V + \varepsilon G_V) \\
& \qquad - (3\sigma_V + \mu_V + \alpha f_T(t)) E_{1V} \\
\dfrac{\mathrm{d}E_{2V}}{\mathrm{d}t} &= 3\sigma_V E_{1V} - (3\sigma_V + \mu_V + \alpha f_T(t)) E_{2V} \\
\dfrac{\mathrm{d}E_{3V}}{\mathrm{d}t} &= 3\sigma_V E_{2V} - (3\sigma_V + \mu_V + \alpha f_T(t)) E_{3V} \\
\dfrac{\mathrm{d}I_V}{\mathrm{d}t} &= 3\sigma_V E_{3V} - (\mu_V + \alpha f_T(t)) I_V \\
\dfrac{\mathrm{d}G_V}{\mathrm{d}t} &= \alpha(1 - f_T(t)) S_V \\
& \qquad - \alpha\Big( f_T(t) + (1 - f_T(t)) p_V \varepsilon \big( \sum_i f_i \dfrac{(I_{1Hi} + I_{2Hi})}{N_{Hi}} + f_A \dfrac{I_A}{N_A} \big) G_V \big) \\
& \qquad\quad - \mu_V G_V
\end{cases} \tag{4}
$$

The actual number of vectors is $S_V, E_{1V}, E_{2V}, E_{3V}, I_V$ and $G_V$ multiplied by $N_V/N_H$, where $N_V$ is the total population of adult tsetse and $N_H = N_{H1} + N_{H4}$ denotes the total human population. Then, the effective probability of human infection per single infective tsetse bite $m_{\text{eff}}$ is defined as $N_V p_H / N_H$ with the original vector-to-human transmission probability $p_H$.

The compartmental ODE model is simulated to compute the disease dynamics in humans, animals and tsetse (see Fig. 2). The total annual passive reported cases for year, $Y$ is calculated by integrating over the new hospitalisations from self-presentation multiplied by the reporting parameter, $u$, to compensate for underreporting of passive cases:

$$P_M = \sum_i \int_Y^{Y+1} \eta_H(T) I_{1Hi}(t) + (\gamma_H(Y) - \gamma_H^{post}(1-u)) I_{2Hi}(t)\,\mathrm{d}t \tag{5}$$

where $i \in$ all human types), whereas the active number of reported cases is given as:

$$\begin{aligned} A_M \quad = \quad & \textstyle\sum_j \text{ proportion screened} \\ & \quad \times \text{ test sensitivity} \\ & \quad\quad \times \text{ compliance} \\ & \quad\quad\quad \times (I_{1Hj}(T) + I_{2Hj}(T)) \\ + \quad & \text{proportion screened} \\ & \quad \times \text{ (1 − test specificity)} \\ & \quad\quad \times \text{ compliance} \\ & \quad\quad\quad \times (N_{Hj}(T) - I_{1Hj}(T) - I_{2Hj}(T)) \end{aligned} \tag{6}$$

where $j \in$ random participants.

**Table 2 Model parameterisation (fixed parameters).** Notation, a brief description, and the values used for fixed parameters.

| Notation | Description | Value | |
|---|---|---|---|
| $N_H$ | Total human population size in 2015 | 41,000 | |
| $\mu_H$ | Natural human mortality rate | 5.1693$\times 10^{-5}$ days$^{-1}$ | [7] |
| $B_H$ | Total human birth rate | $= \mu_H N_H$ | |
| $\sigma_H$ | Human incubation rate | 0.0833 days$^{-1}$ | [8] |
| $\varphi_H$ | Stage 1 to 2 progression rate | 0.0019 days$^{-1}$ | [9, 10] |
| $\omega_H$ | Recovery rate or waning-immunity rate | 0.006 days$^{-1}$ | [11] |
| Sens | Active screening diagnostic sensitivity | 0.952 | [12] |
| $B_V$ | Tsetse birth rate | 0.0505 days$^{-1}$ | [4] |
| $\xi_V$ | Pupal death rate | 0.037 days $^{-1}$ | |
| $K$ | Pupal carrying capacity | $= 111.09 N_H$ | [4] |
| $P$(pupating) | Probability of pupating | 0.75 | |
| $\mu_V$ | Tsetse mortality rate | 0.03 days$^{-1}$ | [8] |
| $\sigma_V$ | Tsetse incubation rate | 0.034 days$^{-1}$ | [13, 14] |
| $\alpha$ | Tsetse bite rate | 0.333 days$^{-1}$ | [15] |
| $p_V$ | Probability of tsetse infection per single infective bite | 0.065 | [8] |
| $\varepsilon$ | Reduced non-teneral susceptibility factor | 0.05 | [2] |
| $f_H$ | Proportion of blood-meals on humans | 0.09 | [16] |
| $\text{disp}_{\text{act}}$ | Overdispersion parameter for active detection | $1 \times 10^{-3}$ | – |
| $\text{disp}_{\text{pass}}$ | Overdispersion parameter for passive detection | $1.5 \times 10^{-4}$ | – |
| $d_{\text{steep}}$ | Speed of improvement in passive detection rate | 10 | Assumed |
| $d_{\text{change}}$ | Switching year for passive improvement | 2014.5 | Assumed |

The value of $B_V$ was chosen to maintain constant population size in the absence of interventions. The steepness and change year of passive detection improvement were selected to yield something close to a step function, with virtually no improvement before the start of 2014 and almost full improvement for 2015.

**Table 3** **Model 4 parameterisation in the new 2000–2019 model fit (fitted parameters).** Notation, brief description, and information on the prior distributions for fitted parameters.

| Notation | Description | Prior distribution[1] | Percentiles of prior distribution [2.5, 50 & 97.5%] | Unit |
|---|---|---|---|---|
| $R_0$ | Basic reproduction number (next generation matrix approach) | $X \sim 1 + \text{Exp}(10)$ | [1.036, 1.053, 1.087] | - |
| $r$ | Relative bites taken on high-risk humans | $X \sim 1 + \Gamma(3.68, 1.09)$ | [2.823, 4.462, 7.635] | - |
| $k_1$ | Proportion of low-risk, random participating people | $X \sim \text{B}(16.97, 3.23)$ | [0.7117, 0.8498, 0.9336] | - |
| $k_4$ | Proportion of high-risk, non-participating people | | $k_4 = 1 - k_1$ | - |
| $\eta_H^{\text{post}}$ | Treatment rate from stage 1, 1998 onwards | $X \sim \Gamma(3.54, 5.32 \times 10^{-5})$ | [9.95, 12.9, 16.4]$\times 10^{-5}$ | days$^{-1}$ |
| $\gamma_H^{\text{post}}$ | Treatment rate from stage 2, 1998 onwards | $X \sim \Gamma(2.45, 0.00192)$ | [4.04, 4.65, 5.36]$\times 10^{-3}$ | days$^{-1}$ |
| $b_{\gamma_H^{\text{pre}}}$ | Relative treatment rate from stage 2 factor, pre-1998 | $X \sim \text{B}(1, 1)$ | [0.799, 0.941, 0.997] | - |
| $\gamma_H^{\text{pre}}$ | Treatment rate from stage 2, pre-1998 | | $\gamma_H^{\text{pre}} = b_{\gamma_H^{\text{pre}}} \gamma_H^{\text{post}}$ | days$^{-1}$ |
| Spec | Active screening diagnostic specificity | $X \sim 0.998 + (1 - 0.998)\,\text{B}(7.23, 2.41)$ | [0.9991, 0.9993, 0.9995] | - |
| $u$ | Proportion of stage 2 passive cases reported | $X \sim \text{B}(20, 40)$ | [0.2082, 0.2612, 0.3254] | - |
| $\eta_{H_{\text{amp}}}$[2] | Relative improvement in passive stage 1 detection rate | $X \sim \Gamma(2.013, 1.049)$ | [0.185, 1.104, 3.093] | - |
| $\gamma_{H_{\text{amp}}}$[2] | Relative improvement in passive stage 2 detection rate | $X \sim \Gamma(1.001, 5)$ | [0.040, 0.525, 8.246] | - |
| $p_{\text{targetdie}}$ | Probability of tsetse hitting a target and dying during host-seeking cycle | $X \sim \text{B}(5.6563, 7.5072)$ | [0.2073, 0.4416, 0.6855] | - |
| S1givenFP | Probability that a false positive cases would be assigned as stage 1 | $X \sim U(0, 1)$ | [0.2588, 0.3901, 0.5444] | - |

[1]Where Exp(.), $\Gamma$(.), B(.) and U(.) are the exponential, gamma (parameterised with shape and scale), beta and uniform distributions, respectively. [2]$\eta_{H_{\text{amp}}}$, $\gamma_{H_{\text{amp}}}$ and $p_{\text{targetdie}}$ are only fitted in the full 2000–2019 fit. The passive amplitude parameters are fixed to 1 (i.e. a doubling of the original rate) and $p_{\text{targetdie}}$ is set to 0.4171 for the updated model 2000–2013 fit.

**Table 4 Model 5 parameterisation in the new 2000–2019 model fit (fitted parameters).** Notation, brief description, and information on the prior distributions for fitted parameters.

| Notation | Description | Prior distribution[1] | Percentiles of prior distribution [2.5, 50 & 97.5%] | Unit |
|---|---|---|---|---|
| $R_0$ | Basic reproduction number (next generation matrix approach) | $X \sim 1 + \text{Exp}(10)$ | [1.041, 1.065, 1.105] | - |
| $r$ | Relative bites taken on high-risk humans | $X \sim 1 + \Gamma(3.68, 1.09)$ | [3.263, 5.663, 10.236] | - |
| $k_1$ | Proportion of low-risk, random participating people | $X \sim \text{B}(16.97, 3.23)$ | [0.6353, 0.7581, 0.8828] | - |
| $k_2$ | Proportion of high-risk, random participating people | $X \sim \text{U}(0, 1)$ | [0.0014, 0.0256, 0.0901] | - |
| $k_3$ | Proportion of low-risk, non-participating people | $X \sim \text{U}(0, 1)$ | [0.0020, 0.0530, 0.1950] | - |
| $k_4$ | Proportion of high-risk, non-participating people | | $k_4 = 1 - k_1 - k_2 - k_3$ | - |
| $\eta_H^{\text{post}}$ | Treatment rate from stage 1, 1998 onwards | $X \sim \Gamma\left(3.54, 5.32 \times 10^{-5}\right)$ | [9.56, 12.5, 16.1]$\times 10^{-5}$ | days$^{-1}$ |
| $\gamma_H^{\text{post}}$ | Treatment rate from stage 2, 1998 onwards | $X \sim \Gamma(2.45, 0.00192)$ | [4.00, 4.62, 5.32]$\times 10^{-3}$ | days$^{-1}$ |
| $b_{\gamma_H^{\text{pre}}}$ | Relative treatment rate from stage 2 factor, pre-1998 | $X \sim \text{B}(1, 1)$ | [0.818, 0.953, 0.998] | - |
| $\gamma_H^{\text{pre}}$ | Treatment rate from stage 2, pre-1998 | | $\gamma_H^{\text{pre}} = b_{\gamma_H^{\text{pre}}} \gamma_H^{\text{post}}$ | days$^{-1}$ |
| Spec | Active screening diagnostic specificity | $X \sim 0.998 + (1 - 0.998)\,\text{B}(7.23, 2.41)$ | [0.9991, 0.9993, 0.9995] | - |
| $u$ | Proportion of stage 2 passive cases reported | $X \sim \text{B}(20, 40)$ | [0.2012, 0.2532, 0.3162] | - |
| $\eta_{H_{\text{amp}}}$[2] | Relative improvement in passive stage 1 detection rate | $X \sim \Gamma(2.013, 1.049)$ | [0.189, 1.082, 3.061] | - |
| $\gamma_{H_{\text{amp}}}$[2] | Relative improvement in passive stage 2 detection rate | $X \sim \Gamma(1.001, 5)$ | [0.039, 0.514, 7.366] | - |
| $p_{\text{targetdie}}$ | Probability of tsetse hitting a target and dying during host-seeking cycle | $X \sim \text{B}(5.6563, 7.5072)$ | [0.2169, 0.4401, 0.6917] | - |
| S1givenFP | Probability that a false positive cases would be assigned as stage 1 | $X \sim U(0, 1)$ | [0.2657, 0.3939, 0.5329] | - |

[1]Where Exp(.), $\Gamma$(.), B(.) and U(.) are the exponential, gamma (parameterised with shape and scale), beta and uniform distributions, respectively. [2]$\eta_{H_{\text{amp}}}$, $\gamma_{H_{\text{amp}}}$ and $p_{\text{targetdie}}$ are only fitted in the full 2000–2019 fit. The passive amplitude parameters are fixed to 1 (i.e. a doubling of the original rate) and $p_{\text{targetdie}}$ is set to 0.4171 for the updated model 2000–2013 fit.

**Table 5 Model 7 parameterisation in the new 2000–2019 model fit (fitted parameters).** Notation, brief description, and information on the prior distributions for fitted parameters.

| Notation | Description | Prior distribution[1] | Percentiles of prior distribution [2.5, 50 & 97.5%] | Unit |
|---|---|---|---|---|
| $R_0$ | Basic reproduction number (next generation matrix approach) | $X \sim 1 + \text{Exp}(10)$ | [1.016, 1.041, 1.075] | - |
| $r$ | Relative bites taken on high-risk humans | $X \sim 1 + \Gamma(3.68, 1.09)$ | [2.297, 4.057, 7.356] | - |
| $k_1$ | Proportion of low-risk, random participating people | $X \sim \text{B}(16.97, 3.23)$ | [0.7155, 0.8526, 0.9356] | - |
| $k_4$ | Proportion of high-risk, non-participating people | | $k_4 = 1 - k_1$ | - |
| $k_A$ | Relative population of animal reservoirs ($k_A = N_A/N_H$) | $X \sim \Gamma(1.26, 19.3)$ | [3.44, 27.7, 101.0] | - |
| $f_A$ | Proportion of blood-meals on animal reservoirs | $X \sim U(0, 1)$ | [0.0128, 0.2225, 0.7465] | - |
| $\eta_H^{\text{post}}$ | Treatment rate from stage 1, 1998 onwards | $X \sim \Gamma\big(3.54, 5.32 \times 10^{-5}\big)$ | [10.3, 13.6, 17.6]$\times 10^{-5}$ | days$^{-1}$ |
| $\gamma_H^{\text{post}}$ | Treatment rate from stage 2, 1998 onwards | $X \sim \Gamma(2.45, 0.00192)$ | [4.05, 4.65, 5.36]$\times 10^{-3}$ | days$^{-1}$ |
| $b_{\gamma_H^{\text{pre}}}$ | Relative treatment rate from stage 2 factor, pre-1998 | $X \sim \text{B}(1, 1)$ | [0.767, 0.929, 0.997] | - |
| $\gamma_H^{\text{pre}}$ | Treatment rate from stage 2, pre-1998 | | $\gamma_H^{\text{pre}} = b_{\gamma_H^{\text{pre}}} \gamma_H^{\text{post}}$ | days$^{-1}$ |
| Spec | Active screening diagnostic specificity | $X \sim 0.998 + (1 - 0.998)\,\text{B}(7.23, 2.41)$ | [0.9991, 0.9993, 0.9995] | - |
| $u$ | Proportion of stage 2 passive cases reported | $X \sim \text{B}(20, 40)$ | [0.2155, 0.2744, 0.3474] | - |
| $\eta_{H_{\text{amp}}}$ [2] | Relative improvement in passive stage 1 detection rate | $X \sim \Gamma(2.013, 1.049)$ | [0.201, 1.098, 2.947] | - |
| $\gamma_{H_{\text{amp}}}$ [2] | Relative improvement in passive stage 2 detection rate | $X \sim \Gamma(1.001, 5)$ | [0.039, 0.623, 9.259] | - |
| $p_{\text{targetdie}}$ | Probability of tsetse hitting a target and dying during host-seeking cycle | $X \sim \text{B}(5.6563, 7.5072)$ | [0.2140, 0.4408, 0.7016] | - |
| S1givenFP | Probability that a false positive cases would be assigned as stage 1 | $X \sim U(0, 1)$ | [0.2484, 0.3855, 0.5318] | - |

[1]Where Exp(.), $\Gamma$(.), B(.) and U(.) are the exponential, gamma (parameterised with shape and scale), beta and uniform distributions, respectively. [2]$\eta_{H_{\text{amp}}}$, $\gamma_{H_{\text{amp}}}$ and $p_{\text{targetdie}}$ are only fitted in the full 2000–2019 fit. The passive amplitude parameters are fixed to 1 (i.e. a doubling of the original rate) and $p_{\text{targetdie}}$ is set to 0.4171 for the updated model 2000–2013 fit.

**Table 6 Model 8 parameterisation in the new 2000–2019 model fit (fitted parameters).** Notation, brief description, and information on the prior distributions for fitted parameters.

| Notation | Description | Prior distribution[1] | Percentiles of prior distribution [2.5, 50 & 97.5%] | Unit |
|---|---|---|---|---|
| $R_0$ | Basic reproduction number (next generation matrix approach) | $X \sim 1 + \text{Exp}(10)$ | [1.016, 1.042, 1.092] | - |
| $r$ | Relative bites taken on high-risk humans | $X \sim 1 + \Gamma(3.68, 1.09)$ | [2.776, 5.278, 10.556] | - |
| $k_1$ | Proportion of low-risk, random participating people | $X \sim \text{B}(16.97, 3.23)$ | [0.5975, 0.7462, 0.8736] | - |
| $k_2$ | Proportion of high-risk, random participating people | $X \sim \text{U}(0, 1)$ | [0.0023, 0.0436, 0.1542] | - |
| $k_3$ | Proportion of low-risk, non-participating people | $X \sim \text{U}(0, 1)$ | [0.0024, 0.0642, 0.2048] | - |
| $k_4$ | Proportion of high-risk, non-participating people | | $k_4 = 1 - k_1 - k_2 - k_3$ | - |
| $k_A$ | Relative population of animal reservoirs ($k_A = N_A/N_H$) | $X \sim \Gamma(1.26, 19.3)$ | [4.00, 26.4, 89.9] | - |
| $f_A$ | Proportion of blood-meals on animal reservoirs | $X \sim U(0, 1)$ | [0.0193, 0.3455, 0.8408] | - |
| $\eta_H^{\text{post}}$ | Treatment rate from stage 1, 1998 onwards | $X \sim \Gamma\big(3.54, 5.32 \times 10^{-5}\big)$ | [10.4, 13.8, 18.6]$\times 10^{-5}$ | days$^{-1}$ |
| $\gamma_H^{\text{post}}$ | Treatment rate from stage 2, 1998 onwards | $X \sim \Gamma(2.45, 0.00192)$ | [3.95, 4.60, 5.36]$\times 10^{-3}$ | days$^{-1}$ |
| $b_{\gamma_H^{\text{pre}}}$ | Relative treatment rate from stage 2 factor, pre-1998 | $X \sim \text{B}(1, 1)$ | [0.757, 0.929, 0.997] | - |
| $\gamma_H^{\text{pre}}$ | Treatment rate from stage 2, pre-1998 | | $\gamma_H^{\text{pre}} = b_{\gamma_H^{\text{pre}}} \gamma_H^{\text{post}}$ | days$^{-1}$ |
| Spec | Active screening diagnostic specificity | $X \sim 0.998 + (1 - 0.998)\,\text{B}(7.23, 2.41)$ | [0.9991, 0.9993, 0.9995] | - |
| $u$ | Proportion of stage 2 passive cases reported | $X \sim \text{B}(20, 40)$ | [0.2168, 0.2820, 0.3615] | - |
| $\eta_{H_{\text{amp}}}$[2] | Relative improvement in passive stage 1 detection rate | $X \sim \Gamma(2.013, 1.049)$ | [0.186, 1.113, 3.172] | - |
| $\gamma_{H_{\text{amp}}}$[2] | Relative improvement in passive stage 2 detection rate | $X \sim \Gamma(1.001, 5)$ | [0.053, 0.765, 10.28] | - |
| $p_{\text{targetdie}}$ | Probability of tsetse hitting a target and dying during host-seeking cycle | $X \sim \text{B}(5.6563, 7.5072)$ | [0.2185, 0.4468, 0.7119] | - |
| S1givenFP | Probability that a false positive cases would be assigned as stage 1 | $X \sim U(0, 1)$ | [0.2537, 0.3916, 0.5300] | - |

[1]Where Exp(.), $\Gamma$(.), B(.) and U(.) are the exponential, gamma (parameterised with shape and scale), beta and uniform distributions, respectively. [2]$\eta_{H_{\text{amp}}}$, $\gamma_{H_{\text{amp}}}$ and $p_{\text{targetdie}}$ are only fitted in the full 2000–2019 fit. The passive amplitude parameters are fixed to 1 (i.e. a doubling of the original rate) and $p_{\text{targetdie}}$ is set to 0.4171 for the updated model 2000–2013 fit.

#### 3 Support for different model variants

To examine which model variants were most supported through fitting to the longitudinal data, we utilised the deviance information criterion (DIC),

$$DIC = -2LL(\bar{\theta}) + 4Var(LL(\theta)) \tag{7}$$

which assigns a lower score to models with high posterior mean log-likelihood whilst penalising models with a larger number of parameters [17]. The relative likelihood of model $i$ was computed using,

$$\text{Relative } DIC = \exp\left((DIC_{min} - DIC_i)/2\right) \tag{8}$$

and was used to compare models. This method was the same as used for previous model fitting and selection [1].

| Model | Random participation | | Non-participation | | Animals | Relative DIC |
|---|---|---|---|---|---|---|
| | Low-risk | High-risk | Low-risk | High-risk | | |
| 1 | X | | | | | $< 10^{-8}$ |
| 2 | X | X | | | | $< 10^{-8}$ |
| 3 | X | | X | | | $< 10^{-8}$ |
| 4 | X | | | X | | 0.7071 |
| 5 | X | X | X | X | | 1 |
| 6 | X | | | | X | $< 10^{-8}$ |
| 7 | X | | | X | X | 0.0010 |
| 8 | X | X | X | X | X | 0.0007 |

**Table 7** Different model structures under consideration and their relative DIC scores for the fitting to 2000–2019 data.

We found that the data most support using Models 4 and 5.

##### 3.1 Ensemble model approach

Individual model fits (M1–M8) contributed to the ensemble model according to their relative DIC scores. Within 2,000 posteriors in our ensemble model, there are 828, 1169, 3 and 0 unique posteriors from M4, M5, M7 and M8 respectively. Our ensemble results are based on 500 randomly selected posteriors from the ensemble model.

### 4 Posteriors of fitted parameters

#### 4.1 Posteriors for Mandoul under different fits

Table 8 shows summaries of posterior distributions of all fitted parameters in the two fits of the updated model.

**Table 8 Ensemble posteriors of fitted parameters.** Notation, brief description, and [2.5th, 50th & 97.5th] percentile of posteriors for fitted parameters.

| Notation | Description | 2000–2013 fit | 2000–2019 fit |
|---|---|---|---|
| $R_0$ | Basic reproduction number (next generation matrix approach) | [1.04, 1.07, 1.10] | [1.04, 1.06, 1.10] |
| $r$ | Relative bites taken on high-risk humans | [3.58, 5.63, 9.12] | [2.97, 5.07, 9.41] |
| $k_1$ | Proportion of low-risk, random participating people | [0.74, 0.85, 0.93] | [0.65, 0.80, 0.92] |
| $k_2$ | Proportion of high-risk, random participating people | [0, 0, 0.001] | [0, 0.01, 0.09] |
| $k_3$ | Proportion of low-risk, non-participating people | [0, 0, 0.006] | [0, 0.01, 0.18] |
| $k_4$ | Proportion of high-risk, non-participating people | [0.07, 0.14, 0.25] | [0.06, 0.15, 0.27] |
| $k_A$ | Relative population of animal reservoirs ($k_A = N_A/N_H$) | [1, 1, 1] | [1, 1, 1] |
| $f_A$ | Proportion of blood-meals on animal reservoirs | [0, 0, 0] | [0, 0, 0] |
| $\eta_H^{\text{post}}$ | Treatment rate from stage 1, 1998 onwards (days$^{-1}$) | [9.71, 12.99, 16.76] $\times 10^{-5}$ | [9.83, 12.58, 16.27]$\times 10^{-5}$ |
| $\gamma_H^{\text{post}}$ | Exit rate from stage 2 (treatment or death), 1998 onwards (days$^{-1}$) | [2.92, 3.53, 4.18] $\times 10^{-3}$ | [4.02, 4.63, 5.31]$\times 10^{-3}$ |
| $b_{\gamma_H^{\text{pre}}}$ | Relative exit rate from stage 2 factor, pre-1998 | [0.82, 0.95, 1.00] | [0.81, 0.95, 1.00] |
| $\gamma_H^{\text{pre}}$ | Exit rate from stage 2 (treatment or death), pre-1998 (days$^{-1}$) | [2.67, 3.31, 4.02] $\times 10^{-3}$ | [3.59, 4.33, 5.09]$\times 10^{-3}$ |
| Spec | Active screening diagnostic specificity | [0.9974, 0.9986, 0.9994] | [0.9991, 0.9993, 0.9995] |
| $u$ | Proportion of stage 2 passive cases reported | [0.20, 0.26, 0.33] | [0.20, 0.26, 0.32] |
| $\eta_{H_{\text{amp}}}$ | Relative improvement in passive stage 1 detection rate | 1 (fixed) | [0.18, 1.08, 3.06] |
| $\gamma_{H_{\text{amp}}}$ | Relative improvement in passive stage 2 detection rate | 1 (fixed) | [0.04, 0.52, 7.67] |
| $p_{\text{targetdie}}$ | Probability of tsetse hitting a target and dying during host-seeking cycle | 0.4171 (fixed) | [0.2157, 0.4424, 0.6873] |
| S1givenFP | Probability that a false positive cases would be assigned as stage 1 | 1 (fixed) | [0.27, 0.39, 0.54] |

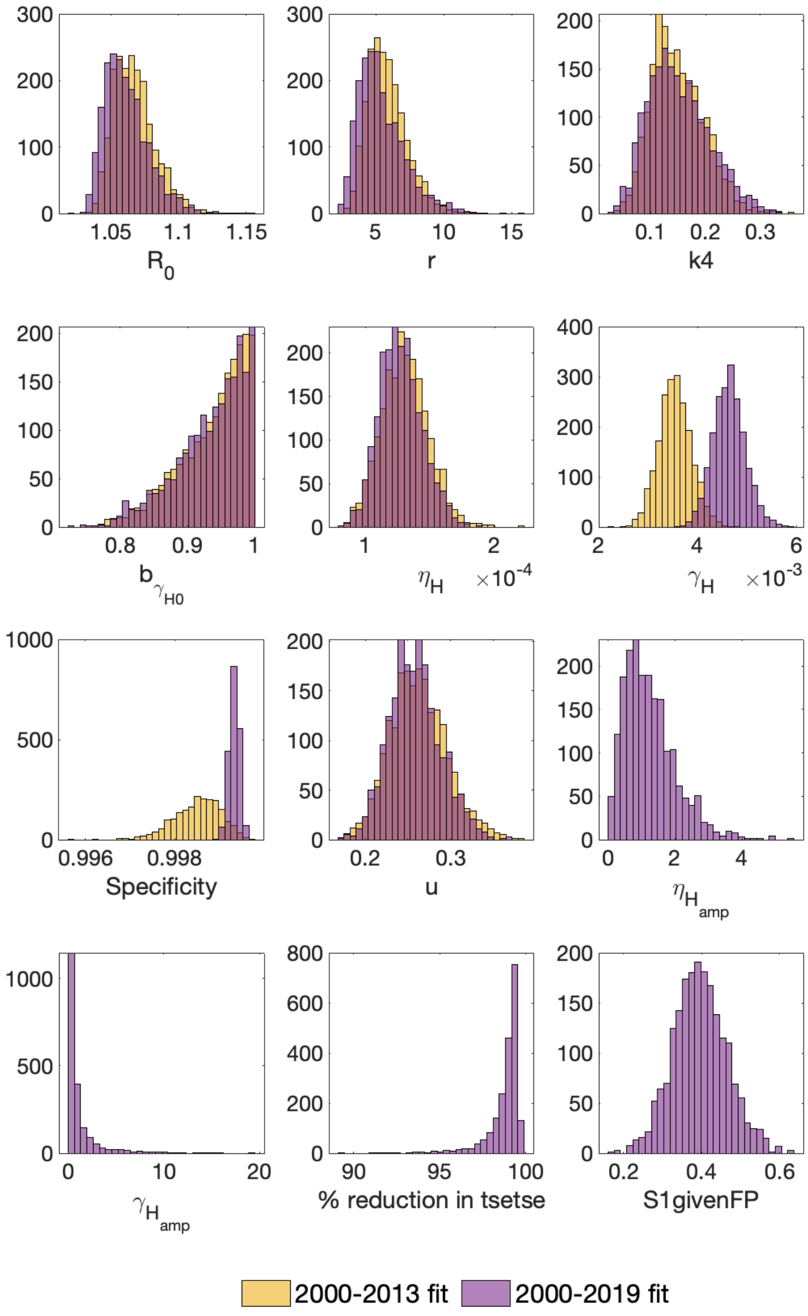


**Figure 3 Posteriors for ensemble models for 2000–2013 fit and 2000–2019 fit.** The histograms show the ensemble model posteriors for the two different fits for each of the fitted parameters. The 2000–2013 fit is shown in yellow and the 2000–2019 fit is shown in red. The 2000–2013 fit did not estimate four of the model parameters which were included in the 2000–2019 fit ($\eta_{H_{\text{amp}}}$, $\gamma_{H_{\text{amp}}}$, $p_{\text{targetdie}}$ and S1givenFP) which were all assumed to be fixed.

#### 5 Additional strategies and cessation criteria

**Table 9** Future strategies (2020 onwards) considered in the present study. AS = active screening, PS = passive screening, VC = vector control.

| Strategy name | AS coverage | PS | VC | Algorithm specificity |
|---|---|---|---|---|
| MaxAS+VC (Imperfect) | Max of 2015–2019 | Continued | Continued | ≈ 99.93% |
| MaxAS+VC | Max of 2000–2019 | Continued | Continued | 100% |

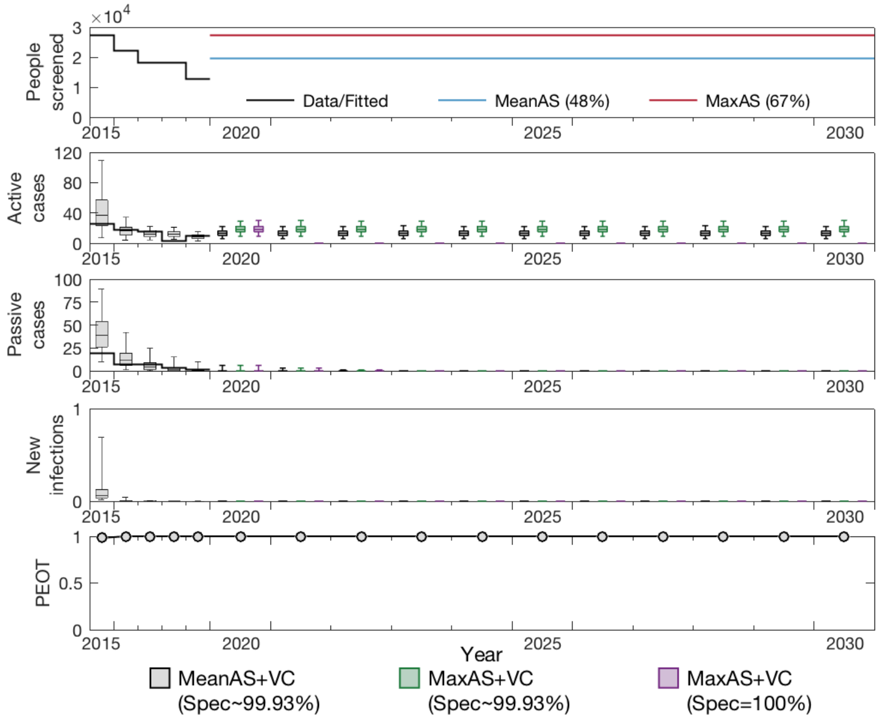


**Figure 4 Projections to 2030.** The ensemble model fitted to data during 2000–2019 was used to make projections under three different strategies. The baseline strategy, MeanAS+VC with imperfect specificity ($\sim 99.93\%$), is denoted by grey boxes. With screening coverage increased to its historical maximum from 2020, the strategy MaxAS+VC with imperfect specificity ($\sim 99.93\%$) is denoted by green boxes. With specificity improved to 100% from 2021, the strategy MaxAS+VC is denoted by purple boxes. All simulations assume PS remains at the level as estimated for 2019 and continues indefinitely. The top panel shows the level of AS assumed in the different projections, the second row shows the active case predictions, the third shows the passive case predictions and the forth shows the expected amount of new infections. The bottom row shows the probability of EOT for each year.

**Table 10** Cessation criteria considered in the present study. AS = active screening.

| Cessation mode | Number of years of zero cases before stopping | Reactive intervention | Number of years of zero cases before stopping reactive intervention |
|---|---|---|---|
| 1 year zero | 1 | AS only | 1 |
| 3 years zeros | 3 | AS only | 1 |
| 5 years zeros | 5 | AS only | 1 |

**Table 11** Summarised statistics for different cessation criteria under two strategies.

| Strategy name | Cessation mode | Cessation probability | Year of cessation [2.5, 50 & 97.5%] | Probability of RAS |
|---|---|---|---|---|
| MeanAS+VC (Imperfect) | 1 year zero | 0 | – | – |
| MeanAS+VC (Imperfect) | 3 years zeros | 0 | – | – |
| MeanAS+VC (Imperfect) | 5 years zeros | 0 | – | – |
| MeanAS+VC | 1 year zero | 1 | [2022, 2022, 2023] | 0.0594 |
| MeanAS+VC | 3 years zeros | 1 | [2024, 2024, 2026] | 0.0086 |
| MeanAS+VC | 5 years zeros | 1 | [2026, 2026, 2028] | 0.0018 |

#### 6 Additional results figures

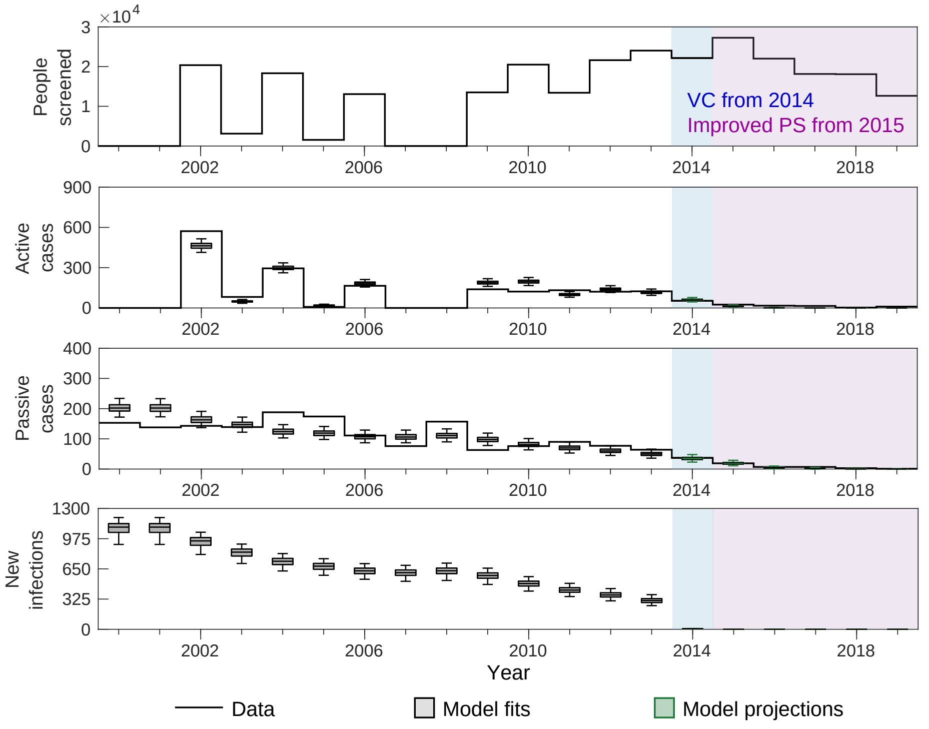


**Figure 5 2000–2013 original ensemble fit.** The top row shows the number of people screened annually in the Mandoul focus from 2000–2019. Shaded regions denote vector control (VC) starting from 2014 (in blue) and improved passive screening (PS) starting from 2015 in purple. The second and third rows show the active and passive case data as a solid black line, with grey-filled box and whisker plots denoting the median (centre line), 50% (box edges) and 95% (whiskers) credible intervals for the original model fit (Mahamat *et al* [1]). Green box and whiskers (2014–2019) show the model projections, updated in the present study to reflect the now known active screening levels, whilst keeping the assumed 99% tsetse reduction from 2014 and doubling of passive detection rates from 2015. Inferred new infections each year are shown on the fourth line.

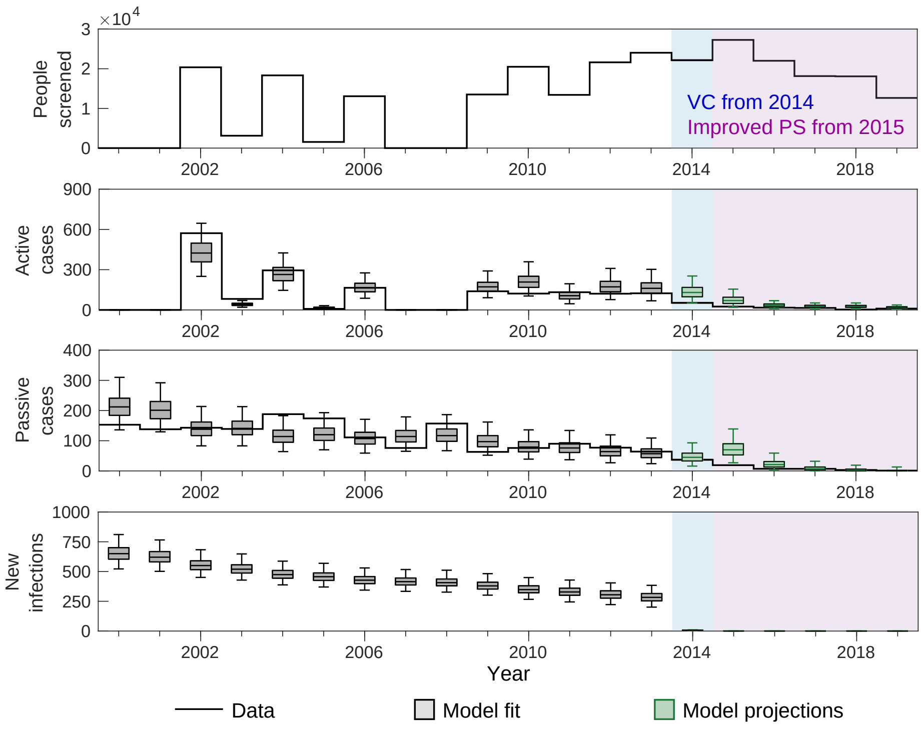


**Figure 6 2000–2013 updated ensemble fit.** The top row shows the number of people screened annually in the Mandoul focus from 2000–2019. Shaded regions denote vector control (VC) starting from 2014 (in blue) and improved passive screening (PS) starting from 2015 in purple. The second and third rows show the active and passive case data as a solid black line, with grey-filled box and whisker plots denoting the median (centre line), 50% (box edges) and 95% (whiskers) credible intervals for the updated model fit using 2000–2013 data only. Green box and whiskers (2014–2019) show the model projections utilising known active screening levels, and assuming a 99% tsetse reduction from 2014 and doubling of passive detection rates from 2015. Inferred new infections each year are shown on the fourth line.

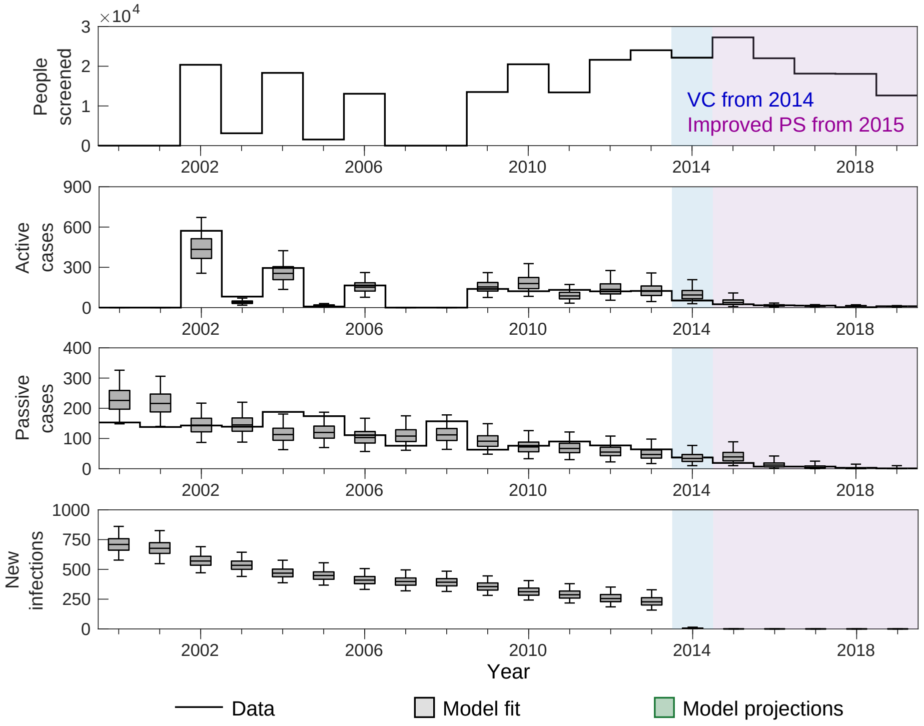


**Figure 7 2000–2019 updated ensemble fit.** The top row shows the number of people screened annually in the Mandoul focus from 2000–2019. Shaded regions denote vector control (VC) starting from 2014 (in blue) and improved passive screening (PS) starting from 2015 in purple. The second and third rows show the active and passive case data as a solid black line, with grey-filled box and whisker plots denoting the median (centre line), 50% (box edges) and 95% (whiskers) credible intervals for the updated model fit using all available case and screening data (2000–2019). The tsetse reduction from 2014 and the passive detection rate improvement from 2015 were fitted. Inferred new infections each year are shown on the fourth line.

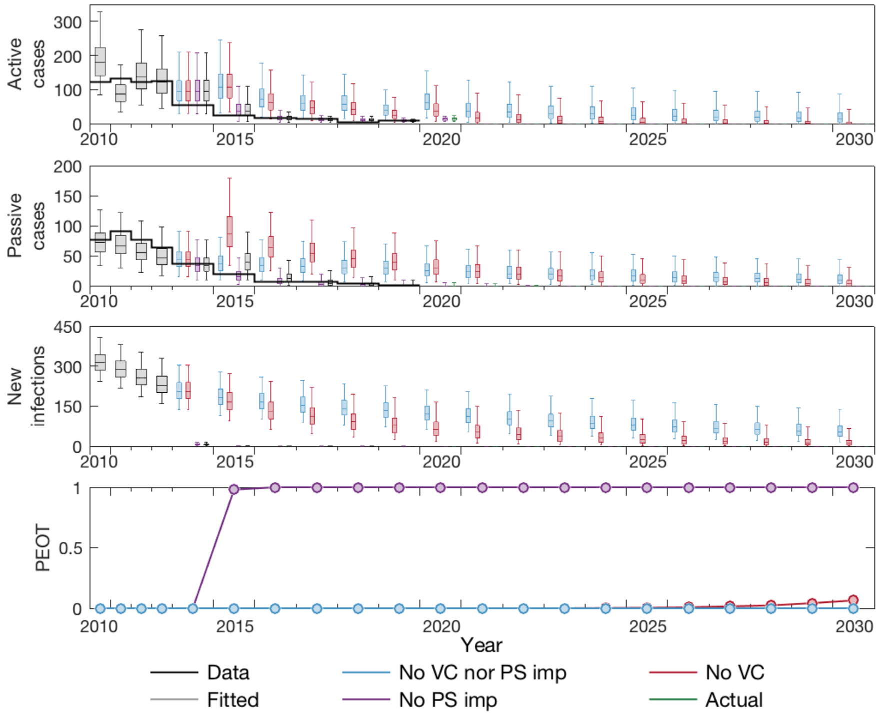


**Figure 8 Counterfactual strategy predictions 2010–2030** The first and second rows show the active and passive case data as a solid black line, with grey-filled box and whisker plots denoting the median (centre line), 50% (box edges) and 95% (whiskers) credible intervals for the updated model fit (2000–2019). Counterfactual scenarios (CFSs) are shown from 2014 in other colours. Blue boxes denote the CFS in which no improvements to either vector control (VC) or passive screening (PS) were made, red boxes denote the CFS in which VC was not deployed but enhanced PS was started in 2015, and purple boxes denote the CFS where VC was deployed in 2014, but no enhanced PS was begun in 2015. From 2020 the projections are run under an assumption of mean active screening. The actual strategy switches from grey to green boxes from that year to reflect that it is a projection rather than fitted. The third row displays the inferred new infections under each scenario, and the last row gives the computed probability of elimination of transmission (EOT) by each year for the different scenarios.

#### 7 Projections with reinvasion of tsetse

To explore the sensitivity of our model system to reinvasion of tsetse after cessation of vector control and possible recrudescence of infection, we have simulated additional scenarios where not only does vector control stop in 2021, but a proportion of the adult tsetse population are also reintroduced to the area. Figure 9A shows the 3 reintroduction scenarios (10%, 50% and 90% of the original adult populations) and the expected bounce-back dynamics of the fly populations following this. Figure 9B then shows the expect outcomes of tsetse bounce-back on new case reporting and new infections.

Ultimately, the extremely low levels of infection estimated to currently be circulating in the human population means that even with the very high (and improbable) levels of reintroduction of 90% tsetse of the 2014 tsetse population, there would be very little additional transmission and we would not expect it would compromise elimination.

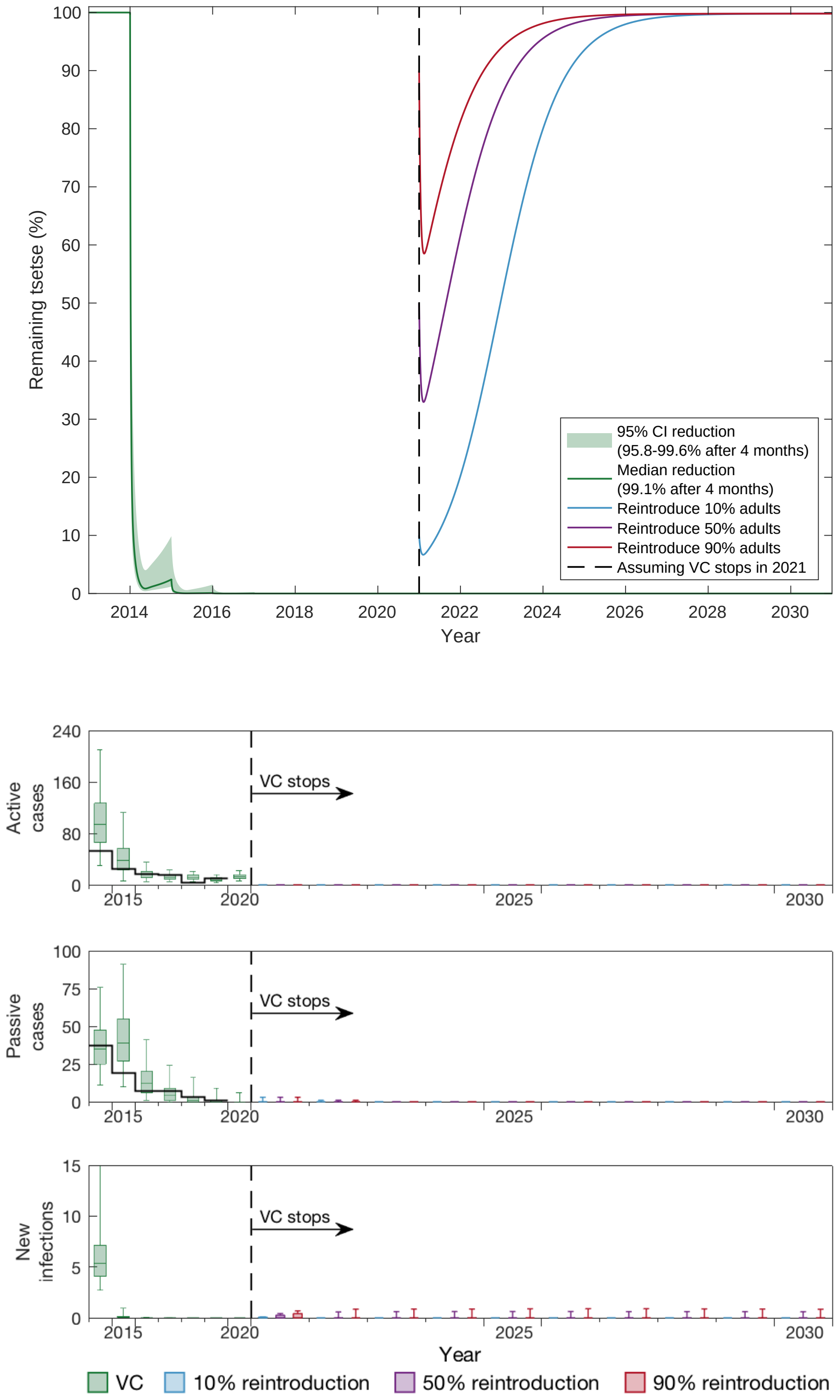


**Figure 9** A. The green line (median) and shaded area (95% CI) shows the estimated VC reduction over 5-year deployment period. We show possible bounce-back scenarios if vector control was stopped in 2021 and either 10%, 50% or 90% of the original adult population was reintroduced. B. Under the three considered bounce-back scenarios we simulate the possible impact on case reporting and new infections until 2030.

#### 8 Serological and parasitological cases

From 2015, there is additional information on how cases were identified as well as staging information. Figure 10 shows the breakdown of cases by those parasitologically confirmed (P+) and those identified through serological tests (S+) or white blood cell count (WBC+). Most (82.1% or 55/67) P+ cases during this time period were identified as stage 2, and in 2019 none of the five P+ cases were stage 1.

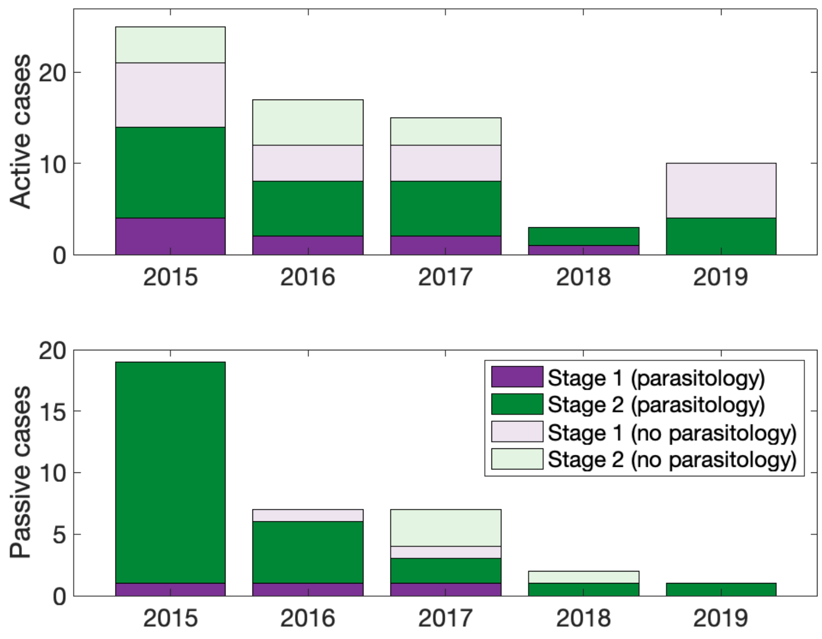


**Figure 10** The stacked bar plots show the proportion of cases in both active (top) and passive (bottom) screening that were confirmed through parasitology (darker colours) or serology or elevated white blood cell count (lighter colours) for the years 2015-2019. Purple bars denote stage 1 and green bars denote stage 2.

#### 9 PRIME-NTD criteria

It has been recommended that good modelling practises should meet the five key principles relating to communication, quality and relevance of analyses - known as Policy-Relevant Items for Reporting Models in Epidemiology of Neglected Tropical Diseases (PRIME-NTD) [18]. We demonstrate how these PRIME-NTD criteria have each been addressed in Table 12.

| | What has been done to satisfy the principle? | Where in the manuscript is this described? |
|---|---|---|
| 1. Stakeholder engagement | This modelling study has been conducted in conjunction with a range of partners, including the national sleeping sickness programme of Chad (PNLTHA-Chad), as well as experts in tsetse control and diagnostics. Many rounds of model fitting and feedback were performed to ensure the modellers factored in critical biology in appropriate ways and simulations produced meaningful outputs which could be used to support appraisal of future strategies. | Authorship list |
| 2. Complete model documentation | Full model fitting code and documentation is available through OpenScienceFramework (OSF). The updated model is fully described in the main text and SI. The previous model is fully described in [1]. | See Materials and Methods section in the main text, Section 2.2 and at OSF (`https://osf.io/rak9d/`) |
| 3. Complete description of data used | The original data and how we aggregated the data for fitting were described in the main text. Aggregate data can be viewed next to model fits in our GUI. | See Materials and Methods section and the GUI (`https://hatmepp.warwick.ac.uk/Mandoulfitandproject/v1/`) |
| 4. Communicating uncertainty | *Structural uncertainty*: The variants of the model selected for the final "ensemble" model were chosen as they had good support compared to other plausible model structures when fitting to the data.<br>*Parameter uncertainty*: We provide estimates for the parameter uncertainty in by showing joint posterior distributions of fitted parameters and providing a summary in the (SI).<br>*Prediction uncertainty*: We represent uncertainty in our results by providing box and whisker plots for fitted dynamics (median, 50% and 95% credible intervals). | *Structural uncertainty*: Methods section in main text and SI.<br>*Parameter uncertainty*: Figures 1 and 2, Figures S3–9 and Table S8<br>*Prediction uncertainty*: Figure 2, Figures S8 and S9 and the GUI (`https://hatmepp.warwick.ac.uk/Mandoulfitandproject/v1/`) |
| 5. Testable model outcomes | First, we used our updated model with censored data (only 2000–2013 for fitting) to test the robustness of the model at predicting the censored case data (i.e. 2014–2019). Second, we used the full model fit (2000–2019) to make future predictions under several plausible strategies, which could be verified in the future as new case data is reported. | See main text results and GUI (`https://hatmepp.warwick.ac.uk/Mandoulfitandproject/v1/`) for validation and future predictions |

**Table 12** PRIME-NTD criteria fulfillment. We summarise how the NTD Modelling Consortium's "5 key principles of good modelling practice" have been met in the present study.

**Author details**
[1]Mathematics Institute, University of Warwick, Academic Loop Road, Coventry, UK. [2]SBIDER, University of Warwick, Academic Loop Road, CV4 7AL Coventry, UK. [3]Independent consultant, Edinburgh, UK. [4]Department of Vector Biology, Liverpool School of Tropical Medicine, Liverpool, UK. [5]Institut de Recherche pour le Développement, UMR INTERTRYP IRD-CIRAD, Université de Montpellier, 34398 Montpellier, France. [6]Swiss Tropical and Public Health Institute, Basel, Switzerland. [7]University of Basel, Basel, Switzerland. [8]Foundation for Innovative New Diagnostics (FIND), Geneva, Switzerland. [9]Programme National de Lutte contre la Trypanosomiase Humaine Africaine (PNLTHA), Moundou, Chad.
